# A Multi-pathogen Hospitalization Forecasting Model for the United States: An Optimized Geo-Hierarchical Ensemble Framework

**DOI:** 10.1101/2025.01.08.25320187

**Authors:** Shaochong Xu, Hongru Du, Ensheng Dong, Xianglong Wang, Liyue Zhang, Lauren M. Gardner

## Abstract

Accurate forecasting of infectious diseases is crucial for timely public health response. Ensemble frameworks have shown promising outcomes in short-term forecasting of COVID-19, among other respiratory viruses, however, there is a need to further improve these frameworks. Here, we propose the Multi-Pathogen Optimized Geo-Hierarchical Ensemble Framework (MPOG-Ensemble), a novel forecasting machine learning framework to forecast state-level hospitalizations of influenza, COVID-19, and RSV in the U.S. This framework is multi-resolution: it integrates state, regionally-trained, and nationally-trained models through an ensemble layer and applies various optimization methods to parameterize the model weights and enhance overall predictive accuracy. This proposed framework builds on existing forecasting literature by 1) employing an ensemble of three spatially hierarchical models with state-level forecasts as the output; 2) incorporating four distinct weight optimization methods to generate the ensemble; 3) utilizing clustering methods to dynamically identify multi-state regions as a function of short-term and long-term hospitalization trends for the regionally-trained model; and 4) providing a generalized multi-pathogen framework to forecast the expected near-term hospitalizations from Influenza, RSV and COVID-19. Results demonstrate MPOG-Ensemble is a robust framework with relatively high performance. Extensive experimentation using historical multi-pathogen data highlights the predictive power of our framework compared to existing ensemble approaches. Its robust performance underscores the framework’s effectiveness and potential for improving and broadening infectious disease forecasting.

## 1. Introduction

The COVID-19 pandemic profoundly altered the landscape of infectious disease epidemiology. Beyond its direct impact on global health, COVID-19 disrupted the established seasonal transmission patterns of other respiratory pathogens such as influenza and respiratory syncytial virus (RSV) ^1–3^. COVID-19 initially led to a decline in other respiratory viruses, largely due to widespread public health measures such as mask-wearing, social distancing, and reduced travel, which collectively limited viral transmission ^4^, but following the relaxation of COVID-19 non-pharmaceutical interventions (NPIs), the simultaneous spread of influenza, COVID-19, and RSV severely strained global healthcare systems and their limited resources. In October 2022, RSV hospitalizations in infants under six months hit a record high, while influenza and COVID-19 also saw substantial increases in hospital admissions^5^.

Future “tripledemics” pose significant threats to public health and health care institutions, as these viruses have not established predictable post-pandemic seasonal patterns ^6^. This uncertainty, combined with early phase of use of RSV vaccines ^7^, underscores the critical need for reliable models to forecast the impact of multiple pathogens simultaneously, to inform public health planning and resource allocation during the post-pandemic era.

While there are very few studies on multi-pathogen forecasting, previous research has shown potential benefits by demonstrating correlations between COVID-19 and influenza cases, where influenza cases were used to predict COVID-19 trends and vice versa ^8^. Other papers primarily focus on demonstrating associations, such as between influenza vaccination rates and COVID-19 outcomes, without utilizing these associations for forecasting ^9^. These parallel trends suggest that a multi-pathogen forecasting approach could be more informative than single-pathogen models. Additionally, NPIs over the first two years of the COVID-19 pandemic not only suppressed the spread of influenza and RSV infections ^10^ but also led to a lack of recent immune responses against these viruses ^11^. Consequently, the easing of restrictions created an ideal environment for the tripledemic to co-circulate, leading to widespread transmission of COVID-19, influenza and RSV.

Existing studies mainly focus on developing models to forecast the spread of single respiratory pathogens such as influenza, COVID-19, and RSV. These studies have leveraged both mechanistic approaches, including the susceptible-exposed-infected-recovered (SEIR) framework ^12–15^, and data-driven statistical methods, leveraging large, diverse datasets from comprehensive surveillance systems to predict infectious disease outcomes ^16^. Some commonly adopted methods includes time-series modelling ^8,17–20^, machine learning techniques like Gaussian processes, multilayer perceptron (MLP), and long short-term memory (LSTM) networks ^21–27^. Various external data sources have been integrated into these models to enhance predictive accuracy, including search engine data, Google Flu Trends data, as well as weather, population, and social media data ^18–26^. However, there remains a clear gap to extend these methods to account for the interrelated dynamics of multiple pathogens simultaneously. This gap is particularly concerning given the overlapping transmission dynamics and symptoms of these viruses, which complicate public health responses and exacerbate the strain on healthcare systems ^28,29^. The simultaneous increases and decreases in emergency department visits for influenza, COVID-19, and RSV observed since October 2022 underscore the critical need for integrated forecasting approaches that can account for the interrelated nature of these pathogens ^5,30^.

In recent years, ensemble models have come to prominence in infectious disease forecasting. These models typically integrate results from different models using specific weighting schemes, such as direct averaging, Expectation-Maximization (EM) selection, and Cluster-Aggregate-Pool approaches ^31,32^. Ensemble methods often outperform single models by potentially reducing prediction errors through decreased variance and, in some cases, bias ^33,34^. A notable example is the CDC’s ensemble model for the FluSight Challenge, which aggregates predictions from multiple teams and has demonstrated exceptional performance across several influenza seasons ^35^. While individual methodologies may not consistently excel, ensemble approaches have shown remarkable reliability in maintaining high performance ^36^. However, optimizing ensemble predictions and determining appropriate model weights remain key challenges in infectious disease forecasting ^31,34,37^. Current ensemble methods typically aggregate results from models at the same geographic level (e.g., county or state), potentially overlooking valuable multi-resolution information. Prior research demonstrated that incorporating trends from different geographic scales could enhance prediction accuracy ^17^. While current ensemble approaches have demonstrated their values, there is potential for further enhancement by incorporating sub-models trained at multiple spatial resolutions and optimizing their integration.

This study builds upon existing infectious disease forecasting literature by introducing the Multi-pathogen Optimized Geo-hierarchical Ensemble Framework (MPOG-Ensemble), a novel approach for forecasting near-term infectious disease hospitalizations for influenza, COVID-19, and RSV simultaneously. MPOG-Ensemble leverages the interconnected dynamics of influenza, COVID-19, and RSV to generate 1-to 4-week state-level hospitalization predictions for all 50 states in the U.S. (See Figure 1 for detail). MPOG-Ensemble is a multi-scale framework that combines spatially explicit sub-models trained across state, regional, and national scales to generate more robust predictions. For the regional sub-model, states are clustered together based on hospitalization trends. The ensemble weights are optimized to minimize recent prediction errors. We evaluate the model performance for all 50 U.S. states from October 14th, 2023, to February 27th, 2024; the MPOG-Ensemble demonstrated increased performance compared to the COVID-19 ForecastHub Ensemble Model and the Flusight Ensemble Model. The model is also being actively submitted to all three hubs ^38–40^ during the 2024-2025 respiratory virus season, highlighting its contribution to real-time forecasting efforts, and ultimately supporting public health decision-making.

**Figure 1.**
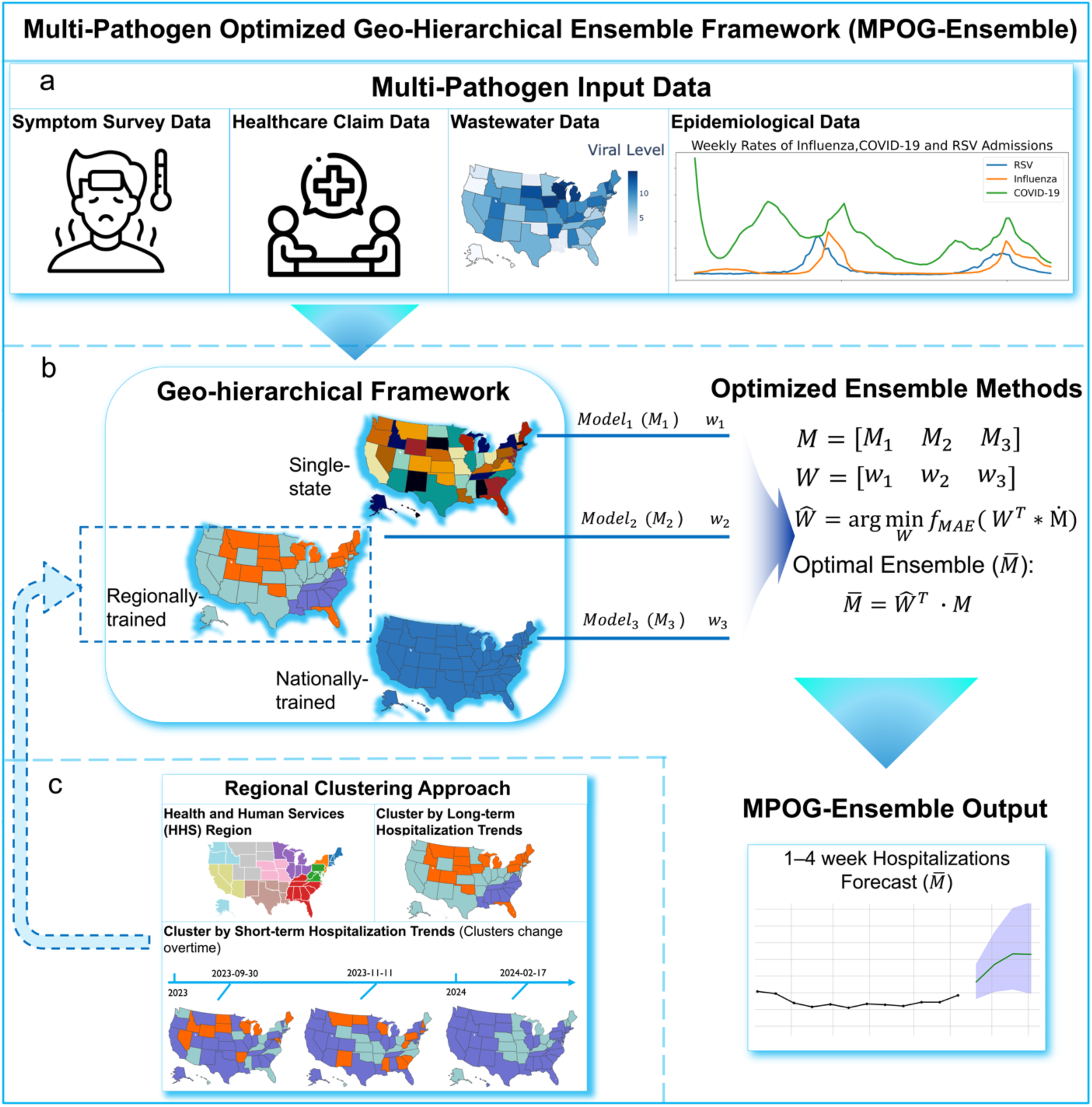
Multi-pathogen Optimized Geohierarchical Ensemble (MPOG-Ensemble) Framework: (a) Input data sources for the framework include symptom survey data, healthcare claim data, wastewater data, and epidemiological data. (b) The main workflow of the framework, which involves training forecasting models at three geospatial levels— single-state, regionally-trained, and nationally-trained — each producing state-level forecasts. These forecasts are then aggregated based on past performance using optimized ensemble methods to generate the final output: a 1-to 4-week hospitalization rate. (c) Regional identification is based on hospitalization rates using Louvain method for the regionally-trained model.

## 2. Data

To effectively train our data-driven model, we incorporated multiple reliable data sources, each offering unique insights into the dynamics of respiratory diseases. Our data selection spans four key categories: 1) symptom survey data derived from google search volume, which serve as a proxy for public concerns and an early indicator for potential outbreaks; 2) healthcare claim data, capturing real-time trends in outpatient visits related to COVID-19 and influenza, offering critical insights into the healthcare system’s response to these illnesses; 3) epidemiological data, which includes traditional epidemiological surveillance data relevant to respiratory diseases; and 4), specifically for COVID-19, wastewater data, which provides an early warning of community-level infections ^41^. All input data are structured as time series with weekly temporal resolution at the state level, with the target variable being state-level hospitalization rates for influenza, COVID-19, and RSV. A detailed description of each included feature is presented in Table 1 below.

**Table 1.** Overview of Variables Considered in the Framework.

| Variable | Description | Source |
| --- | --- | --- |
| Symptom Survey Data derived from Google search volume |  |  |
| Cough-Related Symptoms | The variable is the average Google search volume for key respiratory symptoms—cough, phlegm, sputum, and upper respiratory tract infections—normalized for overall search users. | 42 |
| Nasal Symptoms | This variable represents the average Google search volume, normalized for overall users, for symptoms including nasal congestion, postnasal drip, rhinorrhea, sinusitis, rhinitis, and the common cold. |  |
| Fever Symptoms | Fever, hyperthermia, chills, shivering, and low-grade fever are the symptoms encompassed by this variable, representing the average Google search volume normalized for overall search users. |  |
| Breathing-Related Symptoms | This variable refers to the average Google search volume for symptoms like shortness of breath, wheezing, croup, pneumonia, asthma, crackles, acute bronchitis and bronchitis, normalized for overall search users. |  |
| Throat Symptoms | This variable represents the average Google search volumes for laryngitis, sore throat, and throat irritation—normalized by the total number of search users. |  |
| Healthcare Claim Data |  |  |
| COVID-19 Symptom Visits | This variable tracks the percentage of outpatient doctor visits primarily about COVID-related symptoms. | 42 |
| COVID-19 Confirmed Visits | This variable refers to the percentage of outpatient doctor visits for confirmed COVID-19 cases. |  |
| Influenza Confirmed Visits | This variable represents the percentage of outpatient doctor visits for confirmed influenza cases. |  |
| Epidemiological Data |  |  |
| Hospitalization Rate | This variable is our target variable, including the hospitalization rate for our target pathogen (Influenza, COVID-19, and RSV). | 43–45 |
| Smoothed Hospitalization Rate | This variable represents a 3-week smoothed version of hospitalization data for the target, applied for data augmentation purposes. |  |
| Hospitalization Growth Rate | This variable is also used for data augmentation, serving as an indicator of changes in the hospitalization rate. |  |
| Wastewater Data |  |  |
| COVID-19 Wastewater Level | This variable quantifies the virus level of SARS-CoV-2 virus in wastewater. (For COVID-19 model only.) | 46 |

The training data spans from August 2022 through the forecast date. Forecasts are generated for the period from October 14th, 2023, to February 24th, 2024, encompassing a total of 20 weeks. The reason for selecting data starting from August 2022 is to capture a return to typical influenza activity patterns following the disruptions caused by the COVID-19 pandemic. During the pandemic, influenza hospitalizations significantly decreased. Therefore, the data from the pandemic period stands out as an anomalous period and could potentially skew predictions (See Supplementary Information Figure S.1).

Influenza data covers 50 states, COVID-19 data spans 46 states, and RSV data is available for 12 states. All data was preprocessed at the state level on a weekly basis, with missing values linearly interpolated to ensure the model accurately reflects current epidemiological trends.

## 3. Methods

This section outlines the development of the MPOG-Ensemble framework, covering the following key components: the development of geographically-specific sub-models, the application of regional clustering techniques to identify regions with similar hospitalization trends, the optimization strategy employed for building the ensemble model, and the model performance evaluation method. As shown in Figure 1, we first prepare the multi-pathogen input data. Next, we identify regions for the regionally-trained model (using the identified regions). The data is then fed into three forecasting models—the single-state, regionally-trained, and nationally-trained models. All three models produce state-level hospitalization rates, which are combined using different weighting strategies to generate the final output.

### 3.1. Geo-Hierarchical Architecture

We propose a multi-scale geo-hierarchical architecture framework to capture a wide range of temporal and spatial patterns in hospitalization rates, from state-specific fluctuations to national trends, while accommodating different approaches to regional definition. The framework incorporates geographically-specific sub-models that forecast state-level hospitalization rates, each trained on data with different subsets of states.

#### 3.1.1. Single-State Models

We first implement a hospitalization forecasting model for each state independently, namely the Holt-Winters additive^47^ model for single-state forecasting. The Holt-Winters model is a univariate time series forecasting method that uses only historical hospitalization rates as input (See Supplementary Information Section 2.1. for detail). This model is chosen because of the limited amount of state-specific data, which precludes the use of more complex deep learning models. Compared to Vector Autoregression (VAR) and other multivariate methods, this approach offers simplicity, lower computational demands, and robust performance.

#### 3.1.2. Regionally-trained Models

Geographic clustering can enhance the model’s ability to capture regional variations in disease dynamics, which are often influenced by local factors such as healthcare infrastructure, population density, and public health policies ^48^. Therefore, we employ a regionally-trained model to forecast state-level hospitalizations for groups of states. Because assigning states to regions is subjective, we implement multiple approaches to group states into regions. One widely used regional classification in public health initiatives is the U.S. Department of Health and Human Services (HHS) regions ^49^, but this regional classification is administrative and may not be reflective of regional disease trends. Furthermore, some HHS regions contain as few as two states, potentially providing insufficient data for robust data-driven models and leading to unpredictable results based on historical data ^50^. To address this limitation, we group states with similar hospitalization trends into a set of regions. This approach builds on previous research grouping individual geographic units (e.g., county, state or country) based on similarity in historical influenza-related emergency department visits using methods such as Euclidean Distance (ED) or Dynamic Time Warping (DTW), which has shown promise in predictive capabilities ^51–54^.

Specifically, we apply three clustering algorithms to identify different regions based on hospitalization trends, and then train separate sub-models using the states included in each identified region. By grouping states with similar hospitalization trends, we can generate more accurate and regionally tailored predictions, and mitigate the noise in data that may arise from heterogeneous patterns across different states, allowing the model to focus on meaningful trends and relationships. Specifically, we explored three approaches:

1. **Using HHS regions**: Directly use predefined HHS regions to define state groups for both COVID-19 and influenza. HHS regions are suitable for grouping as public health decisions, interventions, and resource allocations, such as vaccine campaigns, are often implemented using these predefined regions ^17^.
2. **Short-term hospitalization trends clustering**: We employ the Louvain method ^55^, a robust community detection algorithm widely used in network science to generate groups of states to form regions based on recent hospitalization trends. The Louvain method clusters nodes based on the strength and patterns of connections between them, making it highly effective for our purposes. In our network, each state is treated as a node, and the edges between nodes are determined by the correlation between the hospitalization rates timeseries. This correlation measures the similarity in hospitalization trends between states over time. The Louvain method iteratively maximizes a modularity function by merging nodes into same community, which quantifies the density of connections within clusters compared to the random expected connections (See Supplementary Information Section 2.3 and 3.1. for detail). To capture short-term trends, we then cluster states based on short-term hospitalization trends (6 weeks) for COVID-19 and influenza (independently), using a rolling window approach. For each prediction week, we calculate correlations based on the past six weeks’ hospitalization rates, allowing us to identify states with similar trends as they evolve over time. Since different states exhibit varying hospitalization patterns at the same time, the clusters dynamically adjust each week based on the latest hospitalization trends. The six-week period was chosen as it offers an optimal balance between capturing recent shifts in transmission dynamics and smoothing out daily fluctuations.
3. **Long-term hospitalization trends clustering**: We also cluster states based on long-term (seasonal) hospitalization trends for each pathogen to capture more robust patterns over extended periods. For influenza, we create clusters before the first prediction week based on trends from the previous influenza season. These clusters remain fixed and are used consistently throughout the entire forecasting timeframe. For COVID-19, we apply a similar approach by clustering states based on hospitalization trends observed during a key period, specifically the Omicron wave, spanning from December 2021 to September 2023, which marks the beginning of our training phase. This clustering remains fixed across the timeframe, allowing us to use the established patterns for all subsequent predictions.

For our regional level models, we implement multivariate Long Short-Term Memory (LSTM)^27^ networks (See Supplementary Information Section 2.2. for detail). LSTMs have demonstrated robust performance in time series forecasting tasks, particularly when dealing with multiple input variables and complex temporal dependencies. Regional clusters were not generated for RSV due to the limited scope of the RSV-NET, which includes only 12 states.

#### 3.1.3. Nationally-trained Models

For COVID-19 and influenza the national level model utilizes data from all states in a LSTM framework, similar to the regional model (See Supplementary Information Section 2.2. for detail). The nationally-trained RSV model is limited to 12 states for which data is available in RSV-NET.

### 3.2. Optimized Ensemble Methods

Ensemble modeling generates more robust predictions by combining multiple model predictions into one output through model averaging or weighting scheme. Our proposed ensemble model includes the single state model, a regionally-trained model, and the nationally-trained model. The specific regional model is chosen to maximize performance. The models are ensembled using one of four possible weighting schemes based on mean absolute error (MAE), each of which are outlines below.

#### 3.2.1 Ensemble Weighting Methods

The four weighting schemes are 1) direct averaging based on mean absolute error (MEAN), 2) MAE reciprocal weighting (RECI), 3) horizon-weighted MAE reciprocal weighting (RECI by horizon), and 4) linear programming (LP) based on past MAE. Overall, this approach results in 12 distinct ensemble models for COVID-19 and influenza, and 4 models for RSV.

1. For the Direct Averaging (MEAN), we weight the single-state model, regionally-trained models, and nationally-trained models equally.
2. The second method is to optimize the ensemble forecast using RECI, the process begins with calculating the MAE for each model over the past four weeks across multiple prediction horizons. Next, the reciprocals of MAEs are then normalized so that their sum equals one, ensuring a balanced combination of model predictions. Finally, the forecasts are combined using these normalized weights, allowing the ensemble to dynamically adjust based on recent model performance. This approach offers several advantages over simple averaging: First, it emphasizes recent performance, gives higher weight to more accurate models. Moreover, it enhances the ensemble’s sensitivity to error, adaptability to changing conditions, and overall prediction granularity, resulting in more precise and reliable forecasts compared to the MEAN.
3. The RECI by horizon method builds on the reciprocal weighting approach by calculating distinct weights for each prediction horizon instead of using a single weight across all horizons. This refinement allows the model to account for varying accuracy across different time horizons, potentially providing a more reasonable weighting scheme for the ensemble forecasts.
4. Lastly, linear programming (LP) is used to determine the optimal weights that minimize the MAE for predictions made in the most recent week. These optimized weights are then applied to the current prediction week. The optimization process involves assigning a weight to each model in the ensemble such that the sum of the weights equals one, ensuring they are non-negative. The objective function minimizes the average MAE across all observations.

Formulas and additional details can be found in Supplementary Information Section 2.4.

#### 3.2.2 Optimal Ensemble Model Selection

The best-performing MPOG-Ensemble model is selected through a two-step process: First, identifying the regionally-trained model with the smallest MAE. Second, the best ensemble model is determined by comparing MAEs across various weighting strategies applied to the three sub-models. This process is conducted separately for influenza, COVID-19, and RSV predictions. More details can be found in Section 4.3.

### 3.3. Performance Evaluation

We use two metrics to evaluate the predicted weekly hospitalization rates for influenza, COVID-19, and RSV across 1 to 4-week forecasts: relative mean absolute error (MAE) and relative weighted interval score (WIS). These metrics allow us to analyze all ensemble models, select the best-performing model based on relative MAE and are official evaluation metrics in the FluSight Challenge ^43^. The MAE assesses the accuracy of the median (50% quantile) prediction, while the WIS measures the overall distributional accuracy (see Supplementary Information Section 2.5. for details). Smaller values for both metrics indicate better performance. We then compare our model to official published models from the CDC. For influenza forecasts, these include the FluSight Baseline, a median model during the challenge, and the FluSight Ensemble, a well-established model for influenza predictions. For COVID-19 forecasts, we compare our model against the COVIDhub_CDC_baseline (referred to as the COVIDhub Baseline) and the COVIDhub_CDC-ensemble (referred to as the COVIDhub Ensemble). A relative MAE or WIS of 1 indicates baseline-level performance, with values below 1 signifying that our model outperforms the hub baseline model. Further details on our model evaluation are presented in Section 4.2. The relative performance for RSV will not be discussed, as no official ensemble model was available during the prediction period.

Additionally, we conduct a one-tailed Wilcoxon Signed-Rank Test to determine if our prediction results (MAE/WIS) are significantly better than the hub ensemble model. The Wilcoxon Signed-Rank Test is a non-parametric statistical test that compares paired samples to assess whether the median difference between them is significantly different from zero. In this context, the test evaluates whether our model’s prediction errors (MAE/WIS) are consistently lower than those of the hub ensemble model, providing statistical evidence of superior performance. A p-value less than 0.05 would indicate that our model significantly outperforms the hub ensemble model.

## 4. Results

In this section, we present the results of our MPOG-Ensemble framework for 1 to 4-week forecasting of influenza, COVID-19, and RSV at the state level for the United States. Notably, in COVID-19 and FluSight ForecastHub, these 1-to 4-week prediction weeks are represented as 0-3 prediction horizons. First, we present the predictions generated by the optimal MPOG-Ensemble models for selected states to illustrate the model’s forecasting capabilities. Next, we benchmark the overall performance of the MPOG-Ensemble against published CDC models, providing a comparative assessment of its accuracy. To explore the impact of model complexity on performance, we analyze the individual contributions of region clustering and optimized weighting to the ensemble’s predictive strength. Finally, we illustrate how the importance of different geo-hierarchical models varies throughout the evaluation period. For brevity we only present results for influenza below, and present the equivalent results for COVID-19 and RSV in the Supplementary Information. Overall, our findings indicate that the MPOG-Ensemble model delivers robust and precise short-term forecasts, underscoring its potential for real-time public health decision-making.

### 4.1. State-Level Hospitalization Predictions from MPOG-Ensemble

As outlined in Method Section 3.2.2, the optimal ensemble model is selected independently but through same process for Influenza, COVID-19, and RSV, resulting in distinct optimal model configurations for each. The optimized ensembles for each pathogen are as follows:

- Influenza: The best-performing MPOG-Ensemble model for influenza incorporates linear programming for ensemble weighting with short-term trend regional clusters, denoted as MPOG-Ensemble-Influenza (LP, short trend).
- COVID-19: The best-performing MPOG-Ensemble model for COVID-19 employs RECI for ensemble weighting with long-term trend regional clustering, denoted as MPOG-Ensemble-COVID (RECI, long trend).
- RSV: The best-performing MPOG-Ensemble model for RSV uses RECI weighting by forecasting horizon for ensemble weighting, denoted as MPOG-Ensemble-RSV (RECI by horizon). Due to data limitations (only 12 reporting states), RSV forecasting only consists of a nationally-trained model with aggregated single-state models.

The following section illustrates the performance of MPOG-Ensemble-Influenza (LP, short trend) against ground truth data and published CDC models.

Figure 2 illustrates the comparison between predictions made by MPOG-Ensemble-Influenza (LP, short trend) and observations for five selected states during the test period. The five states— Georgia, California, Oregon, Tennessee, and Minnesota—were chosen because they represent different regions of the United States and capture a variety of local conditions and disease dynamics. Across all 50 states, the influenza model achieves an average MAE of 0.311, 0.517, 0.659, and 0.918 for prediction horizons 0 through 3, respectively. Correspondingly, the average WIS for these horizons is 0.238, 0.358, 0.445, and 0.584. The figure highlights a decline in model performance as the forecasting window increases. Predictions for 1- and 2-week horizons closely align with observed values, while 3- and 4-week forecasts show some discrepancies, particularly during peak periods.

**Figure 2.**
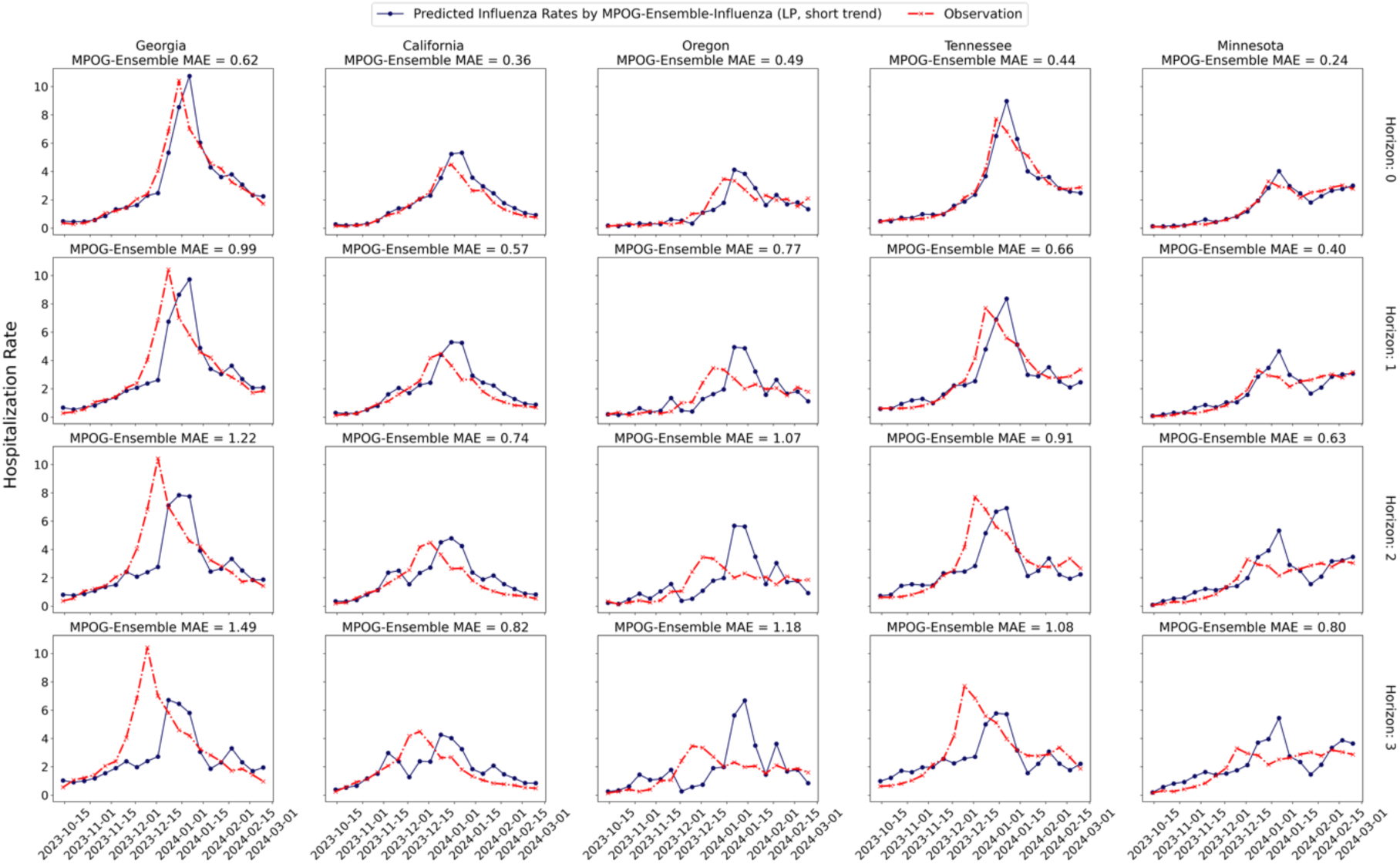
Influenza Hospitalization Predictions for Five Selected States: The dark blue line represents the influenza hospitalization rates predicted by the MPOG-Ensemble-Influenza (LP, short trend), while the red line shows the observed rates during the evaluation period. Each column displays the forecasted and observed rates for a specific state across different prediction horizons, with Horizons 0-3 representing prediction weeks 1-4 as defined by FluSight ForecastHub.

The performance of COVID-19 and RSV forecasts can be found in Supplementary Information Section 3.1

Overall, the MPOG-Ensemble models exhibit strong alignment with the actual targets, particularly during the initial growth and subsequent decline phases. The next section provides a performance comparison between the MPOG-Ensemble and select set of comparative CDC models, offering a comprehensive evaluation of the effectiveness of the proposed framework.

### 4.2. Evaluating the MPOG-Ensemble Models Against Published CDC Hub Models

In this section, we compare the MPOG-Ensemble framework to the set of selected hub models for predicting 1 to 4-week out influenza hospitalization. The comparison for COVID-19 predictions is provided in Supplementary Information Section 3.2. Additionally, we show the statistical significance testing using a one-tailed Wilcoxon Signed-Rank Test to assess whether our model consistently outperforms the hub models across all prediction horizons (in Supplementary Information Section 3.2.1). Our results demonstrate significant improvements in both MAE and WIS, validating the robustness and reliability of our ensemble model in providing accurate short-term hospitalization forecasts for multi-pathogen.

For our influenza comparisons, we selected two models for comparison from the FluSight challenge: the FluSight baseline model, an official CDC model that projects future hospitalization rates by assuming they remain the same as the current week’s rates and outperforms half of the participating models, and the FluSight ensemble model, an official CDC ensemble that combines predictions from all participating teams. Since the FluSight Ensemble ranked in the top 2 among all participating teams during the 2023-2024 influenza season, outperforming it would demonstrate that our model provides superior predictive capabilities compared to a generally robust framework.

As shown in Figure 3, the MPOG-Ensemble-Influenza (LP, short trend) model demonstrates improved performance compared to the FluSight baseline across all forecasting horizons. Specifically, our model shows an average improvement in MAE of 13%, 12%, 15%, and 21% from horizon 0 to horizon 3 compared with FluSight Baseline, respectively. Similarly, the weighted interval score (WIS) also improves by 12%, 15%, 20%, and 24% across the same horizons compared with FluSight Baseline. Our model also surpasses the FluSight Ensemble model, with a 9% improvement in MAE from horizons 0 to 2, a 17% improvement at horizon 3, and an average 5% improvement in WIS. While our model demonstrates strong performance overall, it did not outperform FluSight models in October, suggesting some temporal variability in predictive accuracy. Moreover, as shown in Supplementary Information Table S.2, the results of one-tailed Wilcoxon Signed-Rank test reveal statistically significant improvements in our ensemble model compared to the FluSight Ensemble model across all prediction horizons.

**Figure 3.**
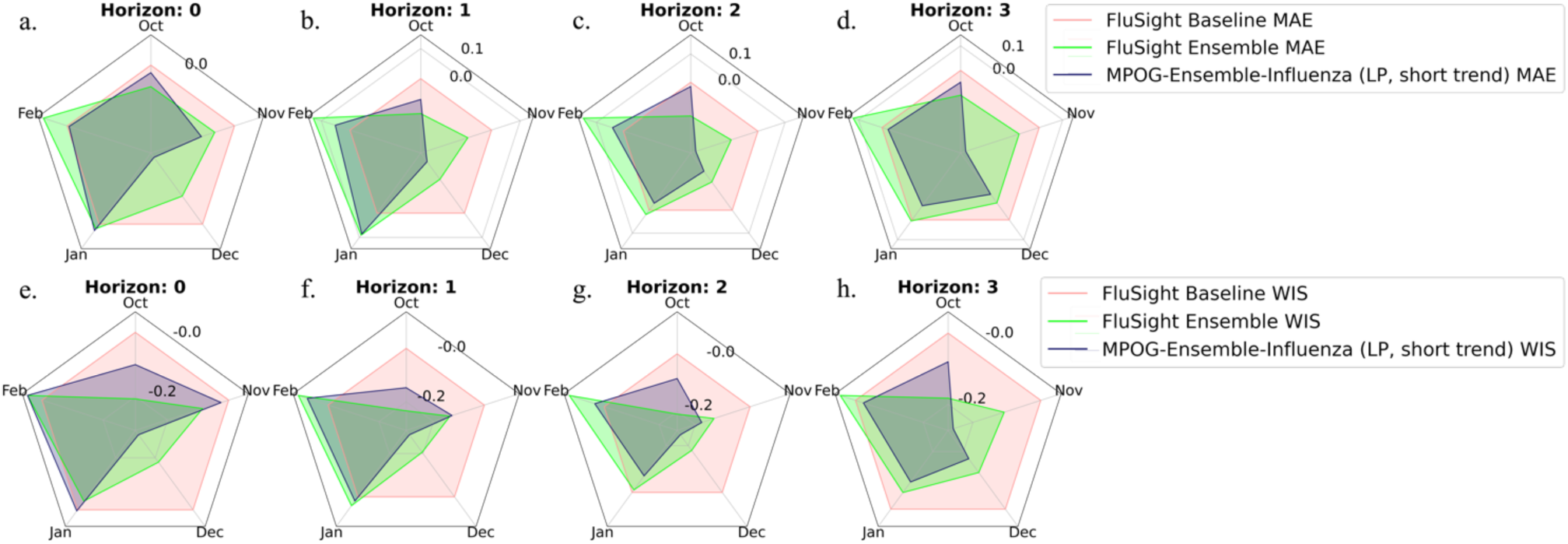
Comparison of Influenza baseline models and MPOG-Ensemble-Influenza (LP, short trend): These figures present radar plots illustrating the median prediction errors across all months for each prediction horizon, where larger magnitudes indicate greater errors. The upper row (a to d) shows the relative MAE compared to the FluSight Baseline Model, while the lower row (e to h) displays the relative WIS. Relative values are calculated by dividing the logscale errors of each model by those of the FluSight Baseline Model. The dark blue line represents the errors of the MPOG-Ensemble-Influenza (LP, short trend), and the light red line corresponds to the FluSight Baseline error, consistently set to 1. The green line indicates the performance of the FluSight Ensemble. A smaller area enclosed within the radar plot reflects a smaller overall error, indicating better performance. The axes are transformed in logarithm scale.

This evaluation of the MPOG-Ensemble-COVID model is detailed in Supplementary Information Section 3.2.

### 4.3. Evaluating Clustering and Ensemble Gains

To further evaluate the contributions of two critical components within our proposed framework, we conduct the follow sensitivity analysis:

1. **Regionalization via Clustering**: We assess the performance of regionally-trained models using different regional definitions, including clusters based on short-term hospitalization trends, clusters derived from long-term trends, and predefined HHS regions.
2. **Optimized Weighting Strategies**: We examine the impact of various optimized weighting strategies within the ensemble model (MEAN, RECI, RECI by horizon, and LP), demonstrating how these approaches enhance performance by adjusting the contributions of individual models.
3. **Dynamic Model Weights**: We evaluate how the weights assigned to the state, regionally-trained, and nationally-trained models change over time, as determined by the optimization methods. We showcase these weight changes specifically for the best-performing models for influenza and COVID-19.

The results are presented in the Supplementary Information Section 3.3. This sub-analysis highlights how increased model complexity enhances performance

## 5. Discussion

In this discussion, we synthesize key findings from our analysis, focusing on how different components of our proposed framework, such as geographic clustering, multi-pathogen integration, and optimized ensemble frameworks, enhance the model performance and stability. Our findings reveal the importance of understanding disease-specific patterns, leveraging interconnected data across pathogens, and applying advanced ensemble techniques to improve decision support during complex, concurrent outbreaks. These insights provide valuable contributions to public health preparedness, highlighting the potential for more effective resource allocation and intervention strategies during respiratory disease seasons.

### 5.1. Multi-pathogen Models Provide Effective Decision Support for Concurrent Outbreaks

The interconnected nature of influenza, COVID-19, and RSV means that data from one pathogen often provide valuable insights into the others ^8^. For example, the symptom survey data we used can be shared across different infectious respiratory diseases, especially during overlapping peak seasons. This interconnected approach enables our framework to make use of shared data sources to improve predictive accuracy for all three pathogens. Specifically, it allows the model to anticipate how a rise in one disease might signal similar trends in others, an aspect particularly important when these illnesses have concurrent peaks that place a strain on healthcare systems. Notably, our forecasts for influenza, COVID-19, and RSV all exhibit high accuracy, achieving small MAE values across all prediction horizons (Figures 2, Supplementary Information Figure S.2, S.3). Compared to traditional pathogen-specific models, such as the FluSight Ensemble and COVIDhub Ensemble, our approach shows improved performance across multiple metrics, including MAE and WIS (Figure 3, Supplementary Information Figure S.4). Another key advantage of using a multi-pathogen model is its ability to provide a comprehensive understanding of the demands on healthcare systems during concurrent outbreaks. When multiple pathogens surge simultaneously, as seen in the past tripledemic, healthcare resources such as hospital beds, staffing, and medical supplies can quickly become overwhelmed. A multi-pathogen model accounts for the overlapping nature of these demands, allowing for more strategic resource allocation that can mitigate the overall burden on the healthcare system.

### 5.2. Geographic Sub-models Enhance Stability and Accuracy of Ensemble Forecasting

This study demonstrates the advantages of using data-driven clustering to define spatial regions for multi-pathogen forecasting. Our results show that models trained on these dynamic regions consistently outperform those based on predefined HHS regions. This improved performance highlights the limitations of relying solely on static regional definitions and underscores the value of capturing localized disease patterns. However, the performance advantage of our data-driven regional models, compared to models trained based on HHS regions, diminished at longer forecasting horizons (2 to 3 weeks), suggesting that longer-term predictions involve more complex dynamics and potentially diverging trends within the initially defined regions. Furthermore, we found that regionally trained models can offer significant advantages over nationally trained models, particularly during periods of shifting hospitalization trends. In these critical phases, regionally trained models demonstrated superior performance and earned greater weight in the ensemble (Supplementary Information Figure S.12). These turning points often represent critical transitions in disease spread (Supplementary Information Figure S.5), where localized factors such as regional disease dynamic and local super spreading events play a significant role ^56^.

The value of combining outputs from three geographic levels (state level, regional level, and national level) to build an ensemble lies in its ability to leverage the strengths of each sub-model, accommodating both localized nuances and broader national trends. By integrating models at different levels, the ensemble achieves more stable performance and outperforms individual sub-models during most periods (Supplementary Information Figure S.9). This stability is crucial during periods of rapid change, such as the onset of a seasonal outbreak or during public health interventions, where different geographic areas may respond differently.

### 5.3. Optimized Ensemble Framework Improves Robustness and Adaptability

Determining which individual model will perform best within an ensemble is inherently challenging, given the variability in disease dynamics and influencing factors ^31^. In this work, we evaluated multiple weighting mechanisms to optimize the ensemble model and found that past performance-based weighting is valuable for enhancing robustness and adaptability, especially in the presence of weaker models. When poorer performing models are part of the ensemble, the linear programming and RECI approach effectively assign lower weights to those models, minimizing their impact on the final prediction (Supplementary Information Figure S.10, S.11). This capability is essential for maintaining forecast performance, particularly during periods of uncertainty, such as the onset of outbreaks or shifts in disease trends. The optimized weighting mechanism ensures that marginal differences in sub-model performance do not degrade the overall ensemble, thereby enhancing the reliability of the forecast.

### 5.4. Influenza Shows Greater Regional Variability Compared to COVID-19

Our analysis highlights the differences in regional patterns between influenza and COVID-19. Influenza exhibits stronger regional trends compared to COVID-19, with peak times and peak sizes varying significantly across different regions. This variability in influenza dynamics is reflected in our clustering results, which reveal more distinct regional patterns for influenza, whereas COVID-19 tends to show a more synchronized trend across states, with similar peak sizes and timing (Supplementary Information Figure S.5, S.6, S.7 and S.8). These findings also manifest in the ensemble model weights, where the influenza ensemble assigns greater weight to single-state models during the increasing phase and regionally-trained models around turning points (Supplementary Information Figure S.12). In contrast, the COVID-19 ensemble shows relatively uniform weighting between models, indicating less regional differentiation (Supplementary Information Figure S.13). These insights suggest that influenza’s regional variation is more influenced by localized factors such as weather conditions, while COVID-19 disease dynamics may be shaped by broader, synchronized influences such as viral characteristics, behavior and public health, consistent with findings from previous studies^57–59^.

### 5.5. Limitations and Future Work

There are some limitations of our study. The ensemble model’s weighting system is based on past performance, which could lead to decreased model performance if a previously high-performing model’s effectiveness declines (especially in light of substantially changing disease dynamics). Additionally, some of the input data streams have become less accessible with time, such as the healthcare claim data and google trends data. Similarly, the incomplete RSV hospitalization data— reported by only 12 states—restricts our ability to develop a meaningful regionally-trained model for RSV forecasting, and thus limits the performance of the RSV model for all states. Such unreliability of public data severely limits the development, evaluation and utility of data-driven models for real-time forecasting, and we strongly encourage better standards and more resources be allocated to support open data for public health modeling.

## 6. Conclusion

The Multi-Pathogen Optimized Geo-Hierarchical Ensemble Framework (MPOG-Ensemble) represents a significant advancement in infectious disease forecasting. By utilizing dynamic clustering for regional modeling, and integrating state, regional, and national level models through an innovative ensemble layer, employing multiple weight optimization methods, MPOG-Ensemble demonstrates superior predictive accuracy for state-level hospitalizations of influenza, COVID-19, and RSV. Our extensive experimentation with historical multi-pathogen data confirms its robust performance. This novel framework not only outperforms existing ensemble approaches but also offers a generalizable solution for multi-pathogen forecasting. The MPOG-Ensemble’s effectiveness highlights its potential to enhance infectious disease forecasts accuracy, thereby supporting more timely and informed public health responses in the face of complex, interrelated respiratory disease dynamics.

## Data Availability

All data produced are available online at Github

https://github.com/Shawn-Tsui/Multi-Pathogen_Optimized_Geo-Hierarchical_Ensemble_Framework.

## CRediT Authorship Contribution Statement

**Shaochong Xu:** Writing – review & editing, Writing – original draft, Software, Methodology, Formal analysis, Visualization, Validation, Data curation, Conceptualization. **Hongru Du:** Writing – review & editing, Methodology, Formal analysis, Visualization, Conceptualization, Investigation, Supervision. **Ensheng Dong:** Writing – review & editing, Visualization, Investigation. **Xianglong Wang:** Data curation. **Liyue Zhang:** Validation. **Lauren M. Gardner:** Writing – review & editing, Supervision, Resources, Funding acquisition, Project administration, Conceptualization.

## Data and Code Availability Statement

The code that implements the framework and the data that support the findings of this study are publicly available in Github repository at https://github.com/Shawn-Tsui/Multi-Pathogen_Optimized_Geo-Hierarchical_Ensemble_Framework.

## Supplementary Information for

### 1. Supplementary Information for Background

Figure S.1 below presents the hospitalization rates for influenza and RSV from 2021 to 2024. It highlights that influenza and RSV admissions were significantly affected by the COVID-19 pandemic, disrupting their typical seasonal patterns and resulting in a notable reduction in cases.

**Figure S.1:**
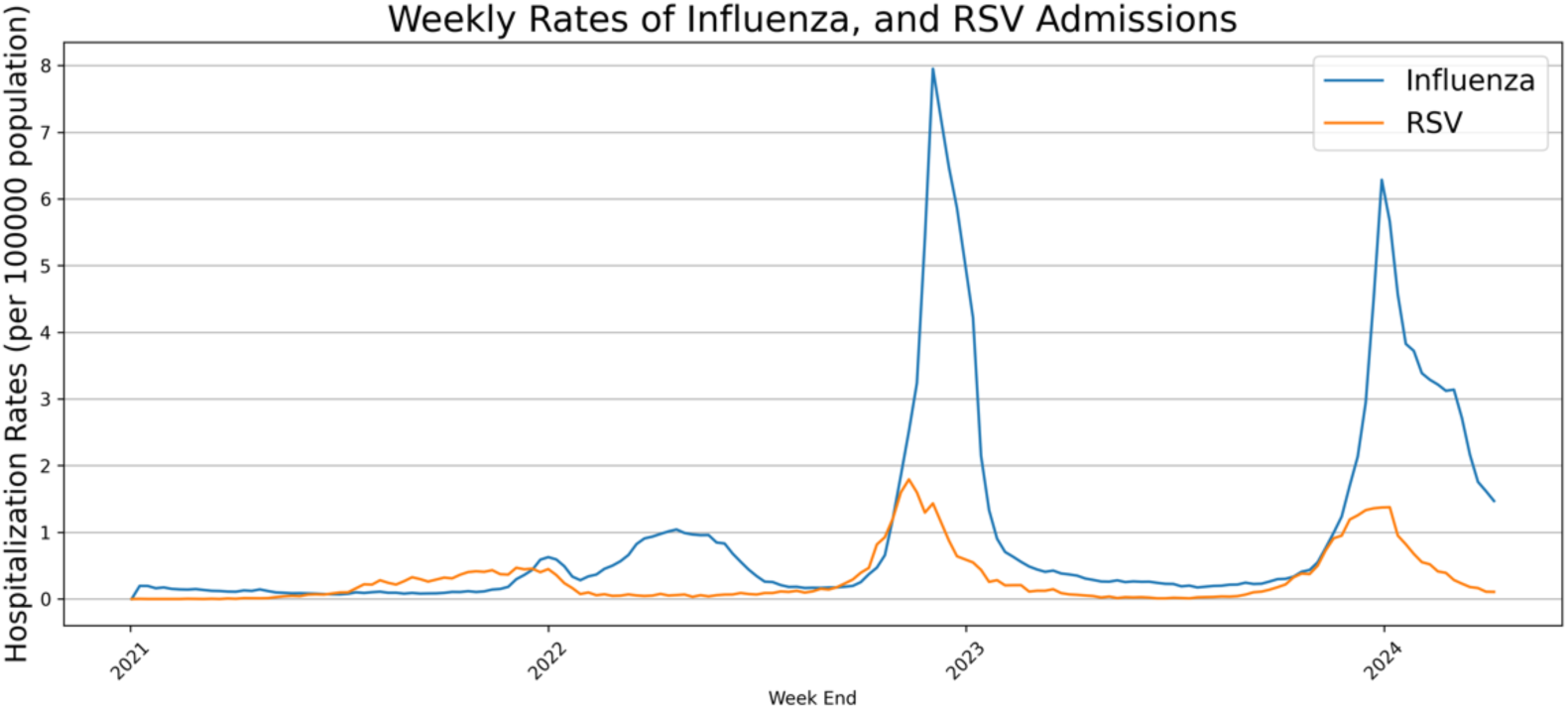
Hospitalization Rates during pandemic: The blue line represents the hospitalization rate for Influenza (per 100,000 population), and the orange line indicates the rate for RSV, covering the period from 2021 to 2024.

### 2. Detailed Explanation of Methods

#### 2.1. Single-state Model

The single-state model is built by Holt-Winters additive method ^1^, it forecasts time series data with both trend and seasonal variations. The model consists of three components: level, trend, and seasonality.

Notations:

*y_t_*: The observed value at time t

*L_t_*: The level (or base value) at time t

*T_t_*: The trend (or slope) at time t

*S_t_*: The seasonal component at time t

*s*: The length of the seasonal cycle

α: Smoothing parameter for the level (between 0 and 1)

β: Smoothing parameter for the trend (between 0 and 1)

γ: Smoothing parameter for the seasonality (between 0 and 1)

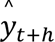: Forecasted value for ℎ periods ahead

Level (*L_t_*):

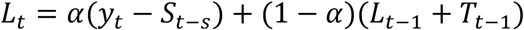

Trend (*T_t_*):

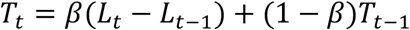

Seasonality (*S_t_*):

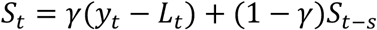

Forecast (*y_t_*):

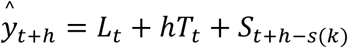

where *k* is the number of complete seasonal cycles between *t* and *t* + *h*.

This is a univariate time series forecasting method. The input for each pathogen model is the time series of hospitalization rates for that specific pathogen (e.g., influenza, RSV, or COVID-19). The model adjusts for trends, seasonality, and fluctuations in hospitalization patterns for each pathogen individually.

#### 2.2. Regionally-trained/ Nationally-trained Model

For both our regionally-trained and nationally-trained models, we employed a Long-Short Term Memory (LSTM) architecture ^2^. The training data was transformed into sequences to predict hospitalization rates per 100,000 people, providing forecasts one to four weeks ahead for each state. To enable probabilistic outputs, we incorporated a dropout layer before the final output layer to introduce prediction variability. Specifically, the model generated predictions 1,000 times, and the forecasted values at specified quantiles were selected to represent the range of uncertainty in the predictions.

The training and validation data spanned from August 2022, with an 80% training and 20% validation split. For real-time forecasting, the test dataset comprised new, unseen data following the prediction date. At each forecast iteration, the training set was updated incrementally by adding one week of newly available data.

We employed the Smoothed L1 loss function, defined as:

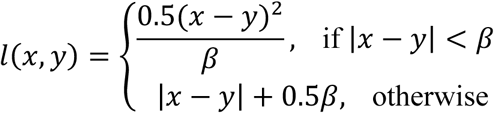

Our models utilized a dropout rate of 0.8 and were trained for 50 epochs with a batch size of 2. Hyperparameter optimization was performed using a grid search across predefined values for learning rate, sequence length, number of layers, and hidden layer size. Early stopping, based on validation error measured by Mean Absolute Error (MAE), was implemented to prevent overfitting.

#### 2.3. Louvain Method

The Louvain Method is used for defining regions before training our regionally-trained model. It is a community detection algorithm that identifies clusters within a network by optimizing modularity, a measure of the strength of division of a network into clusters. In our approach, the correlation of hospitalization rate trends is used as a measure of similarity to construct the network. The method is iterative, starting with each node (state) in its own cluster and then merging clusters to maximize modularity, resulting in defined regions that better reflect local hospitalization patterns and regional variations.

Correlation ρ between two states i and j is calculated as follows:

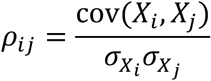

where *X_i_*, *X*_j_ are the time-series data of hospitalization rates for states i and j, respectively, *cov*(*X_i_ X_j_*) is the covariance, and σ_*Xi*_, σ_*Xj*_ are the standard deviations of the time-series data.

Modularity *Q* is defined by:

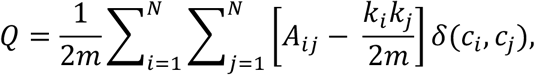

- *A_ij_* represents the the edge weight between nodes *i* and *j*.
- *k_i_* and *k*_*j*_ are the sum of the weights of the edges attached to nodes *i* and *j*, respectively.
- m is the sum of all of the edge weights in the graph.
- N is the total number of nodes in the graph.
- *c_i_* and *c*_*j*_ are the communities to which the nodes *i* and *j* belong.
- δ is Kronecker delta function, δ = 1 if *c*_*i*_ and *c*_*j*_ are the same cluster; otherwise, δ = 0.

The following illustrates the iterative process of the Louvain method:

##### Algorithm 1: Louvain Method^3^

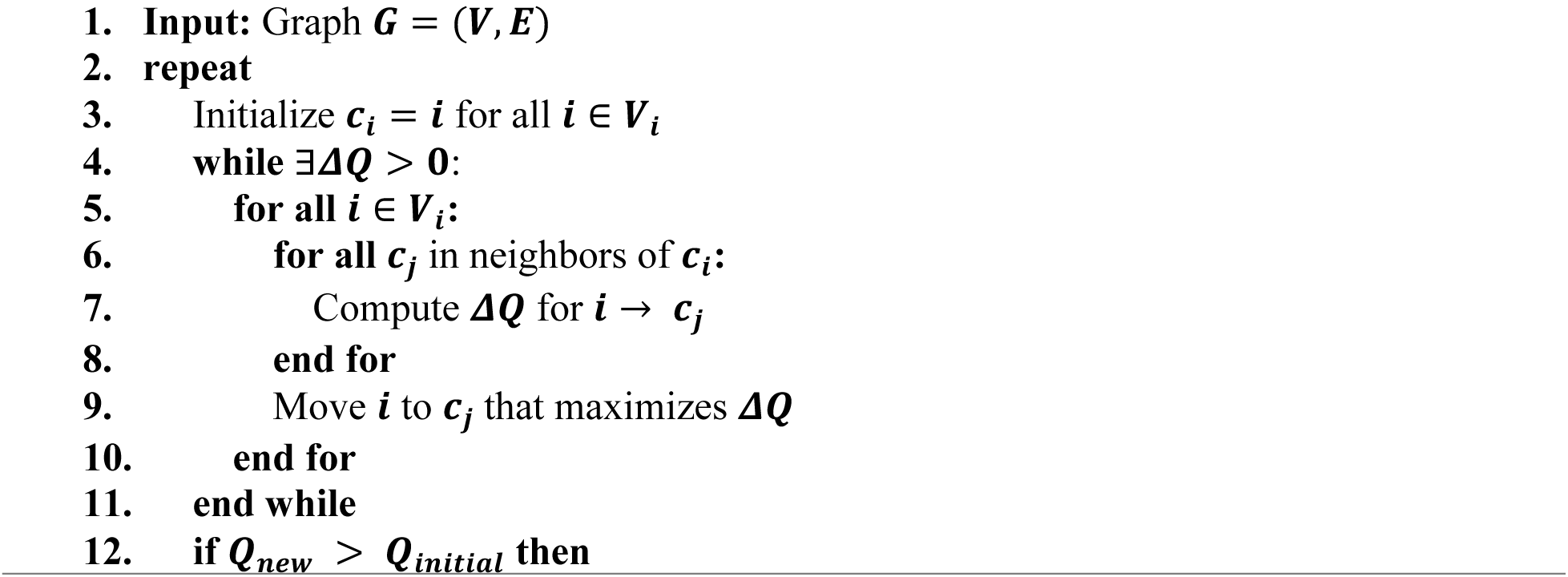
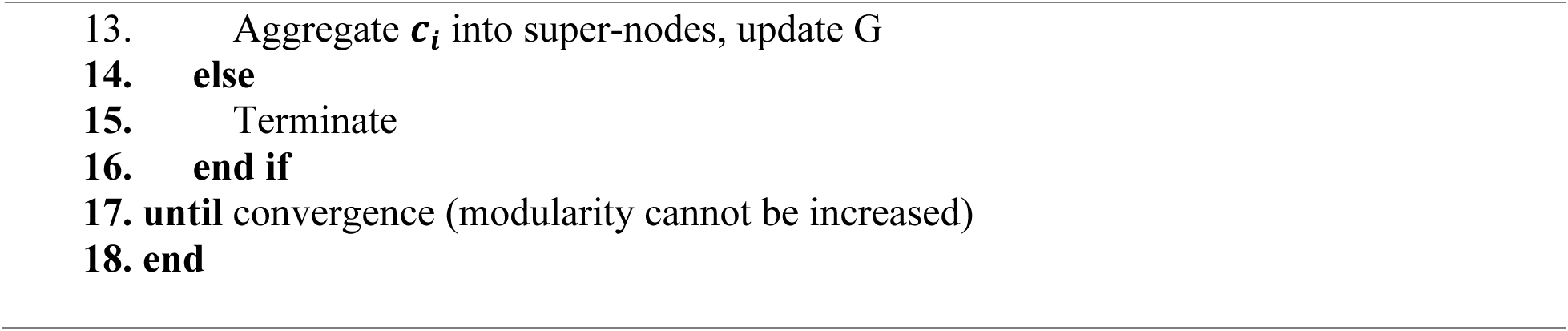

#### 2.4. Optimized Ensemble Methods

In this section, we provide detailed explanations for the steps involved in the methods outlined in Section 3.2. Optimized Ensemble Methods.

##### 2.4.1. MAE reciprocal weighting (RECI)

1. **Calculate MAE**: Suppose at the prediction week t, for each model over the past 4 weeks, we have the following ten MAE based on historical prediction: **Table S.1.**
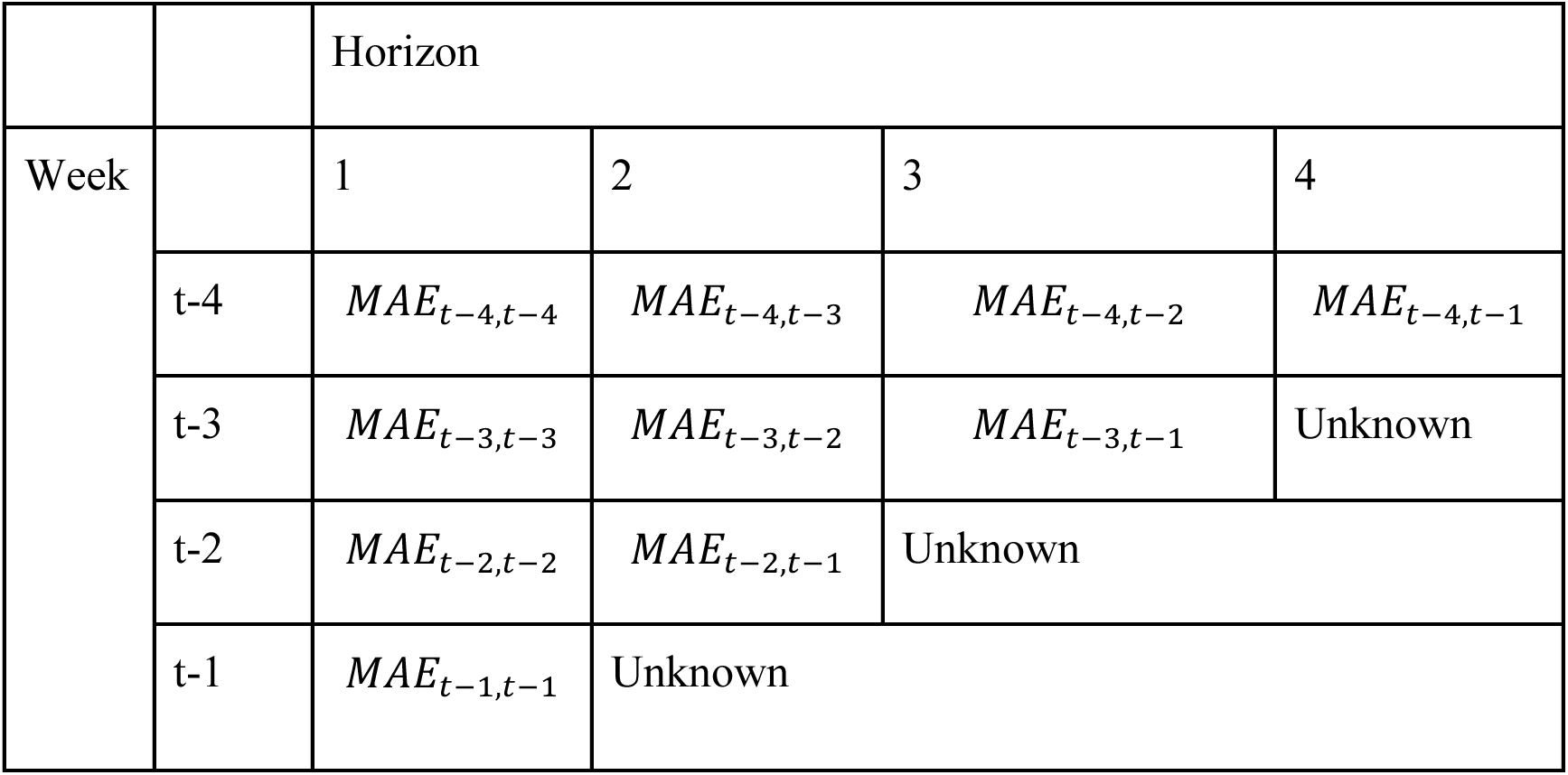
Mean Absolute Error (MAE) Over the Past 4 Weeks for Each Prediction Horizon at Prediction Week t.

and the MAE is calculated by the mean of these ten values

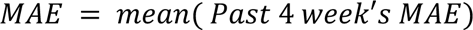

### 2. Compute the Reciprocal of Each MAE and Normalize These Weights

First, calculate the reciprocal of each MAE:

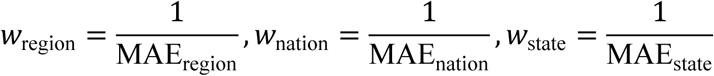

Then, normalize these weights so that their sum is 1:

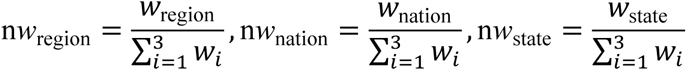

### 3. Combine Forecasts

Finally, combine the forecasts using the normalized weights:

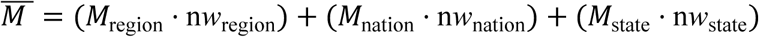

Where 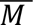 represents the ensemble results and M is the forecasts of each sub-model.

#### 2.4.2. Horizon-weighted MAE reciprocal weighting (RECI by horizon)

This method is analogous to the previous one but calculates weights separately for each prediction horizon. The weight is given by

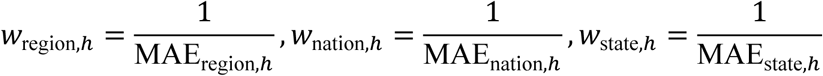

where h represents horizon h.

#### 2.4.3. Linear Programming (LP)

In this section, we formulate the linear programming problem to find the optimal weights that minimize the Mean Absolute Error (MAE) for week *t* – *i_th_*, where i is selected from {1, 2, 3, 4}. These weights are then applied to the prediction for week t. The formulation is given by:

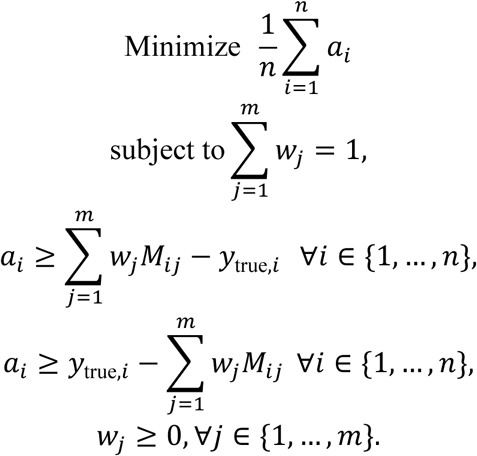

- n: Number of observations.
- m: Number of models in the ensemble.
- *a_i_*: Absolute error for observation i.
- *W_i_*: Weight for model j.
- *M_i_*_-_: Prediction from model j for observation i.
- *y_true,i_*: True value for observation i.

### 2.5. Weighted Interval Score

The quantile score for a single quantile level τ is defined as:

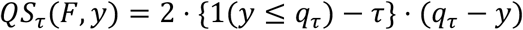

where:

- *q*_τ_ is the τ-quantile of the predictive distribution *F*.
- 1(⋅) is the indicator function, which is 1 if the condition inside is true and 0 otherwise.

The Weighted Interval Score (WIS) ^4^ is calculated as follows:

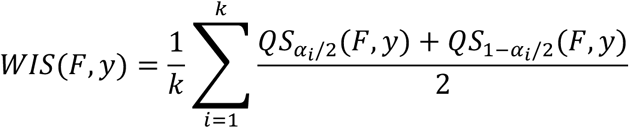

Where:

- *k* is the number of prediction intervals.
- α_*i*_ is the significance level for the *i*-th prediction interval.
- *QS*_Bα*i*/+_(*F*, *y*) is the quantile score for the lower quantile.
- 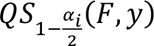 is the quantile score for the upper quantile.

## 3. Supplementary Information for Results

### 3.1. State-Level Hospitalization Predictions from MPOG-Ensemble

**Figure S.2:**
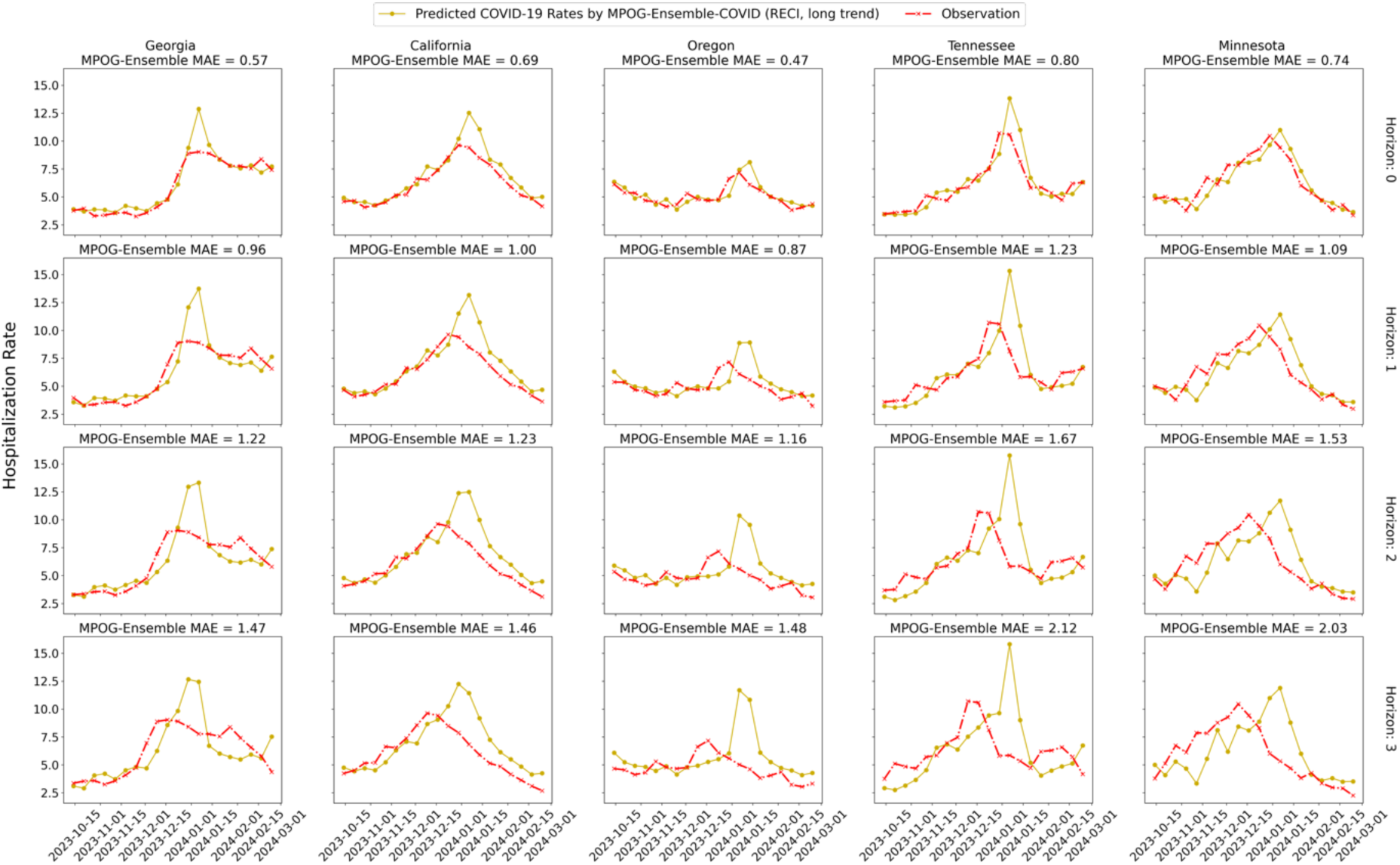
COVID-19 Hospitalization Predictions for Five Selected States: The goldenrod line represents the COVID-19 hospitalization rates predicted by the MPOG-Ensemble-COVID (RECI, long trend), while the red line shows the observed rates during the evaluation period. Each column displays the forecasted and observed rates for a specific state across different prediction horizons, with Horizons 0-3 representing prediction weeks 1-4 as defined by COVID-19 ForecastHub.

Figure S.2 illustrates the comparison between predictions made by MPOG-Ensemble-COVID (RECI, long trend) and observations for five selected states during the test period. Across all states, the influenza model achieves an average MAE of 0.634, 0.905, 1.229, and 1.531 for prediction horizons 0 through 3, respectively. Correspondingly, the average WIS for these horizons is 0.428, 0.584, 0.753, and 0.941. The COVID-19 predictions show similar patterns as influenza with relative larger average MAE and WIS.

**Figure S.3:**
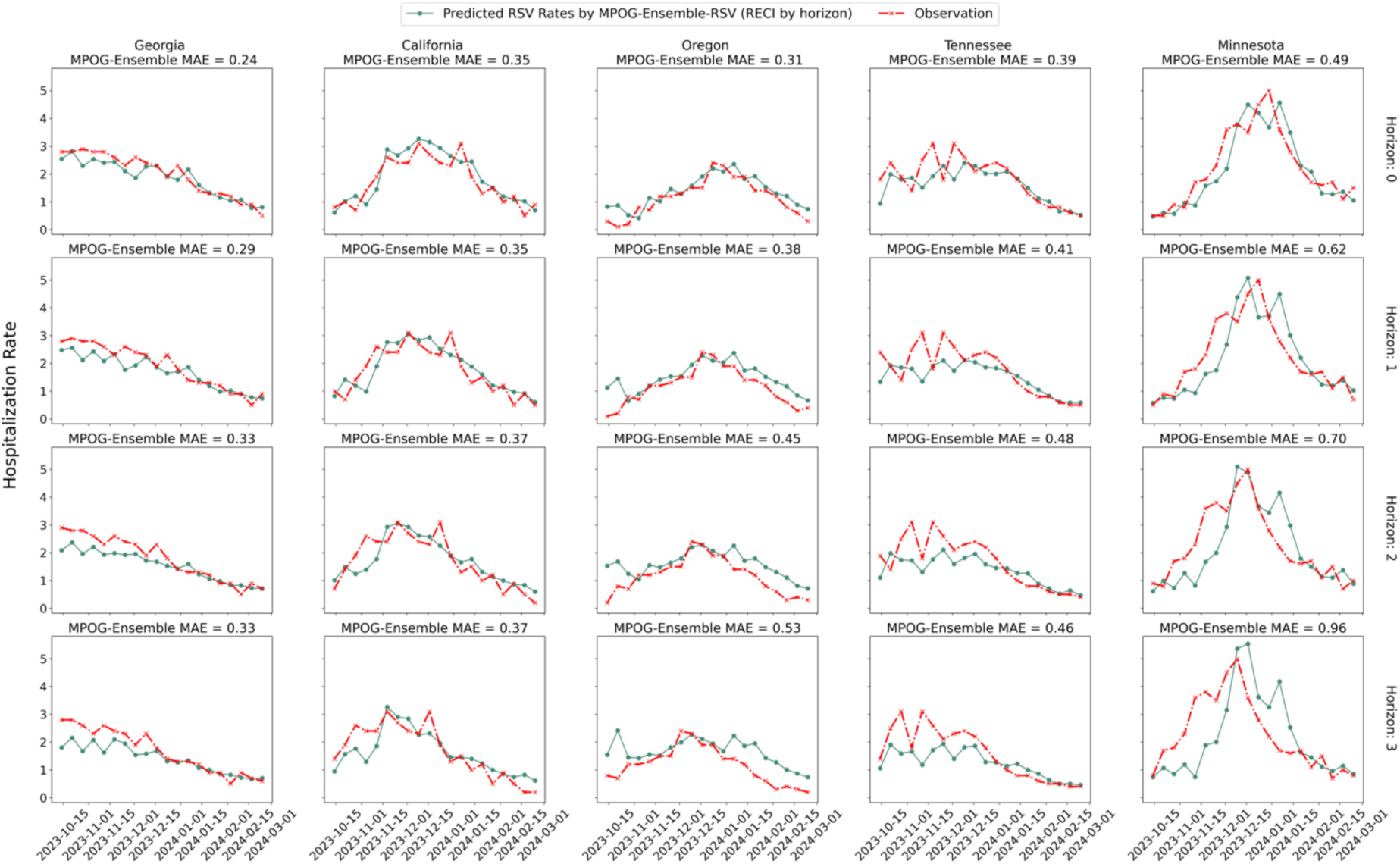
RSV Hospitalization Predictions for Five Selected States: The green line represents the RSV hospitalization rates predicted by the MPOG-Ensemble-RSV (RECI by horizon), while the red line shows the observed rates during the evaluation period. Each column displays the forecasted and observed rates for a specific state across different prediction horizons.

Figure S.3 illustrates the comparison between predictions made by MPOG-Ensemble-RSV (RECI by horizon) and observations for five selected states during the test period. Across all states, the RSV model achieves an average MAE of 0.401, 0.457, 0.516, and 0.599 for prediction horizons 0 through 3, respectively. Correspondingly, the average WIS for these horizons is 0.269, 0.305, 0.341, and 0.386. Compared to influenza and COVID-19 forecasts, the RSV model demonstrates better alignment with observed targets and relatively smaller MAE and WIS values. These results suggest that RSV may be more predictable than influenza and COVID-19.

### 3.2. Evaluating the MPOG-Ensemble Models Against Published CDC Hub Models

#### 3.2.1. One-tailed Wilcoxon Signed-Rank Test

**Table S.2.** **One-tailed Wilcoxon Signed-Rank Test for FluSight Ensemble and MPOG-Ensemble-Influenza (LP, short trend)**

| Horizon | Wilcoxon Signed-Rank Test | *P*-value |
| --- | --- | --- |
| 0 | MAE | $P < .001$ |
| 1 | MAE | $P < .001$ |
| 2 | MAE | $P < .001$ |
| 3 | MAE | $P < .001$ |
| Overall | MAE | $P < .001$ |
| 0 | WIS | $P < .001$ |
| 1 | WIS | $P < .001$ |
| 2 | WIS | $P < .001$ |
| 3 | WIS | $P < .001$ |
| Overall | WIS | $P < .001$ |

In the one-tailed Wilcoxon Signed-Rank Test, we hypothesize that the MAE and WIS of the FluSight Ensemble model are greater than those of our ensemble results. The resulting p-values indicate a statistically significant improvement in our ensemble model across all prediction horizons, confirming its improved performance.

**Table S.3.** One-tailed Wilcoxon Signed-Rank Test for COVIDhub Ensemble model and MPOG-Ensemble-COVID (RECI, long trend)

| Horizon | Wilcoxon Signed-Rank Test | $P$-value |
| --- | --- | --- |
| 0 | MAE | $P < .001$ |
| 1 | MAE | $P < .001$ |
| 2 | MAE | $P < .001$ |
| 3 | MAE | $P < .001$ |
| Overall | MAE | $P < .001$ |
| 0 | WIS | $P < .001$ |
| 1 | WIS | $P < .001$ |
| 2 | WIS | $P < .001$ |
| 3 | WIS | $P < .001$ |
| Overall | WIS | $P < .001$ |

In our evaluation of COVID-19 forecasting performance, our proposed ensemble model demonstrates a statistically significant improvement in accuracy across all forecast horizons compared to the COVIDhub Ensemble baseline. The Wilcoxon Signed-Rank Test yielded p-values less than 0.001 for each forecast horizon, providing robust evidence to reject the null hypothesis of equal performance. This consistent superiority across prediction horizons suggests that MPOG-Ensemble-COVID (RECI, long trend) offers a substantial advancement in predictive capability.

#### 3.2.2. Comparison of MPOG-Ensemble Models with Published CDC Hub Models

For COVID-19 forecasting comparisons, we use two baseline models: the COVIDhub_CDC-ensemble model and the COVIDhub_CDC-baseline model. These will be referred to as the COVIDhub Ensemble and COVIDhub Baseline, respectively. These models are similar in structure and purpose to the corresponding models from FluSight.

**Figure S.4:**
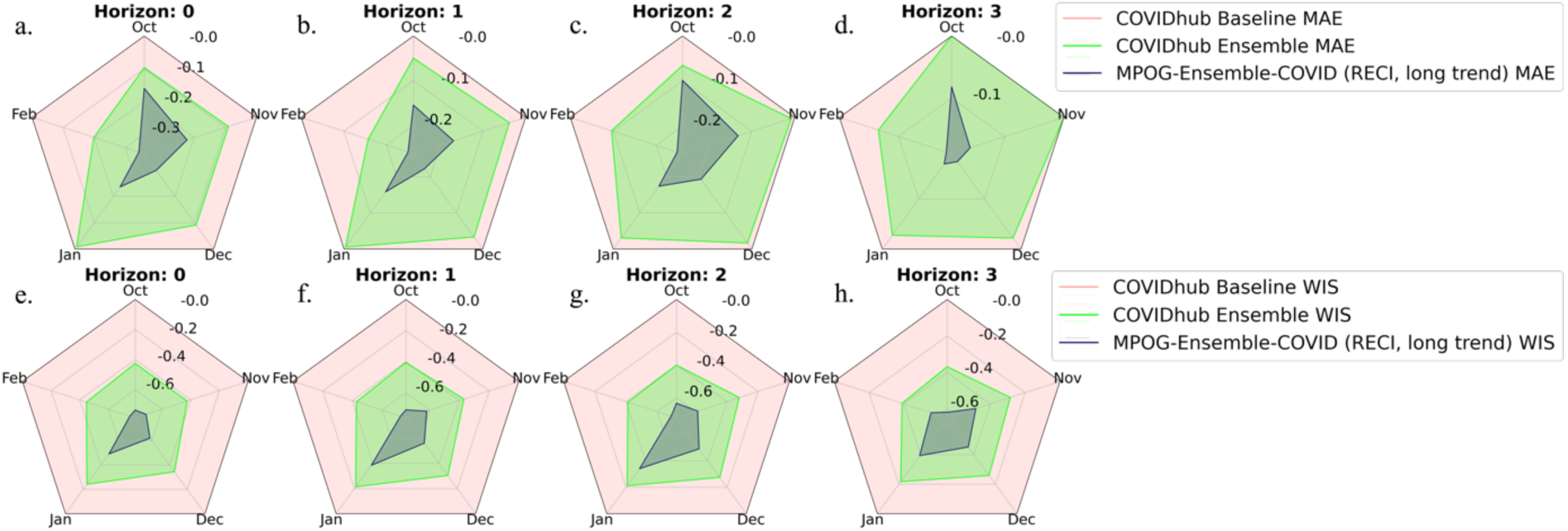
Comparison of COVID-19 baseline models and MPOG-Ensemble-COVID (RECI, long trend): the figures in the upper row display the relative MAE, while the lower row presents the relative WIS. These values are calculated by dividing the true value by the COVIDhub Baseline model. The radar plot illustrates the median error across all months for each prediction horizon, where a larger magnitude indicates a larger error. The dark blue line shows the error from our MPOG-Ensemble-COVID (RECI, long trend), and the light red line represents the COVIDhub Baseline error, which is consistently set to 1. The green line indicates the performance of the COVIDhub Ensemble. A smaller area on the radar plot corresponds to a smaller error, demonstrating better performance. The axes are transformed in logarithm scale.

Figure S.4 reveals that our model consistently outperforms both the COVIDhub Baseline and COVIDhub Ensemble models across all months and forecast horizons, as measured by MAE and WIS. When compared specifically to the COVIDhub Ensemble model, our approach demonstrates substantial improvements in accuracy. For MAE, we observe reductions of 30%, 27%, 24%, and 26% for forecast horizons 1 through 4, respectively. The improvements in WIS are more pronounced, with reductions of 48%, 44%, 40%, and 36% across the horizons. These significant margins of improvement underscore the improved predictive capabilities of our model compared to the CDC’s ensemble approach. As shown in Supplementary Information Table S.3, the one-tailed Wilcoxon Signed-Rank test results further confirm that our ensemble model achieves statistically significant improvements over the COVIDhub Ensemble model across all prediction horizons.

The results for RSV will not be discussed in other sections, as the lack of sufficient data prevents the development of a meaningful regionally-trained model.

### 3.3. Evaluating Clustering and Ensemble Gains

#### 3.3.1 Regionalization via Clustering

Our short-term trend clustering method generates dynamic regions that evolve with changing influenza hospitalization rates (Figure S.5). This approach captures localized patterns by allowing the number of regions and their state memberships to shift over time. To illustrate this dynamic, we examine three representative weeks: early November (early season), early January (peak), and February (rapid decline). For instance, in November 2023, Mississippi and Georgia were clustered into Region 3, both experiencing an initial decrease in hospitalization rates followed by a subsequent increase. However, by January 2024, their trajectories diverged. Mississippi, with a continuous rise in hospitalization rates, was placed in Region 2, while Georgia, where rates began to decline, was assigned to Region 1. Finally, in February 2024, both states were clustered back into the same region, reflecting a shared pattern of decline following the peak. Figure 8 visually demonstrates these shifts, highlighting how our method adapts to the evolving trends. This dynamic clustering provides a more accurate representation of evolving local patterns compared to static regionalization methods.

**Figure S.5:**
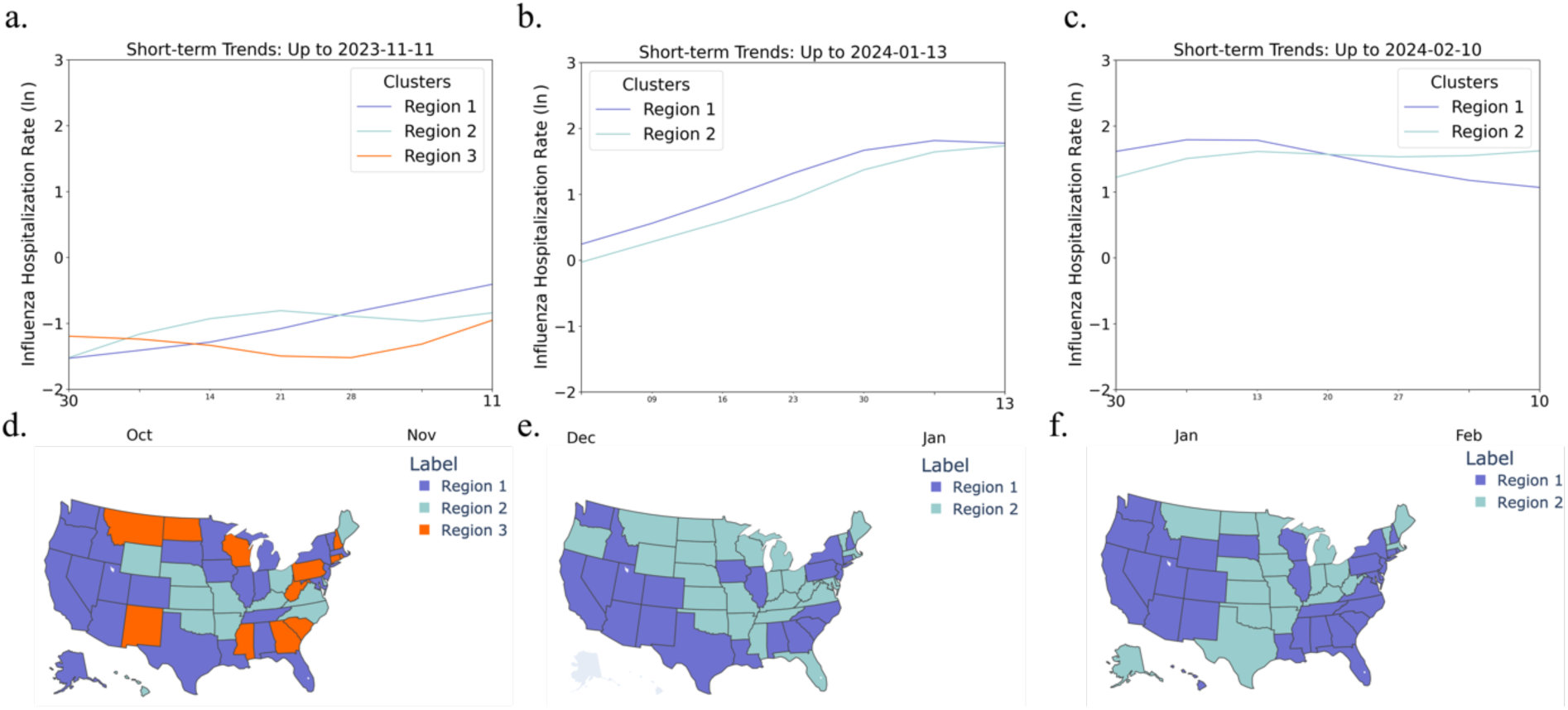
Short-term Hospitalization Trends Clustering for Influenza Hospitalization Rate in Selected Prediction Weeks: Subfigures (a, b, c) display the smoothed median influenza hospitalization rates for each clustered region (transformed using the natural logarithm, ln), while subfigures (d, e, f) show the corresponding states within those regions. The number of regions and the states within each region change dynamically based on short-term hospitalization trends.

Figure S.6 shows the clustering results for COVID-19 hospitalization rates during three selected prediction weeks, highlighting how the number of regions and the states within each region evolve over time due to changes in hospitalization patterns. For instance, New Hampshire and Maine belong to different regions on ‘2023-11-11’ and ‘2024-01-13’ but are grouped in the same region on ‘2024-02-10’, reflecting how shifting hospitalization trends influence regional clustering.

**Figure S.6:**
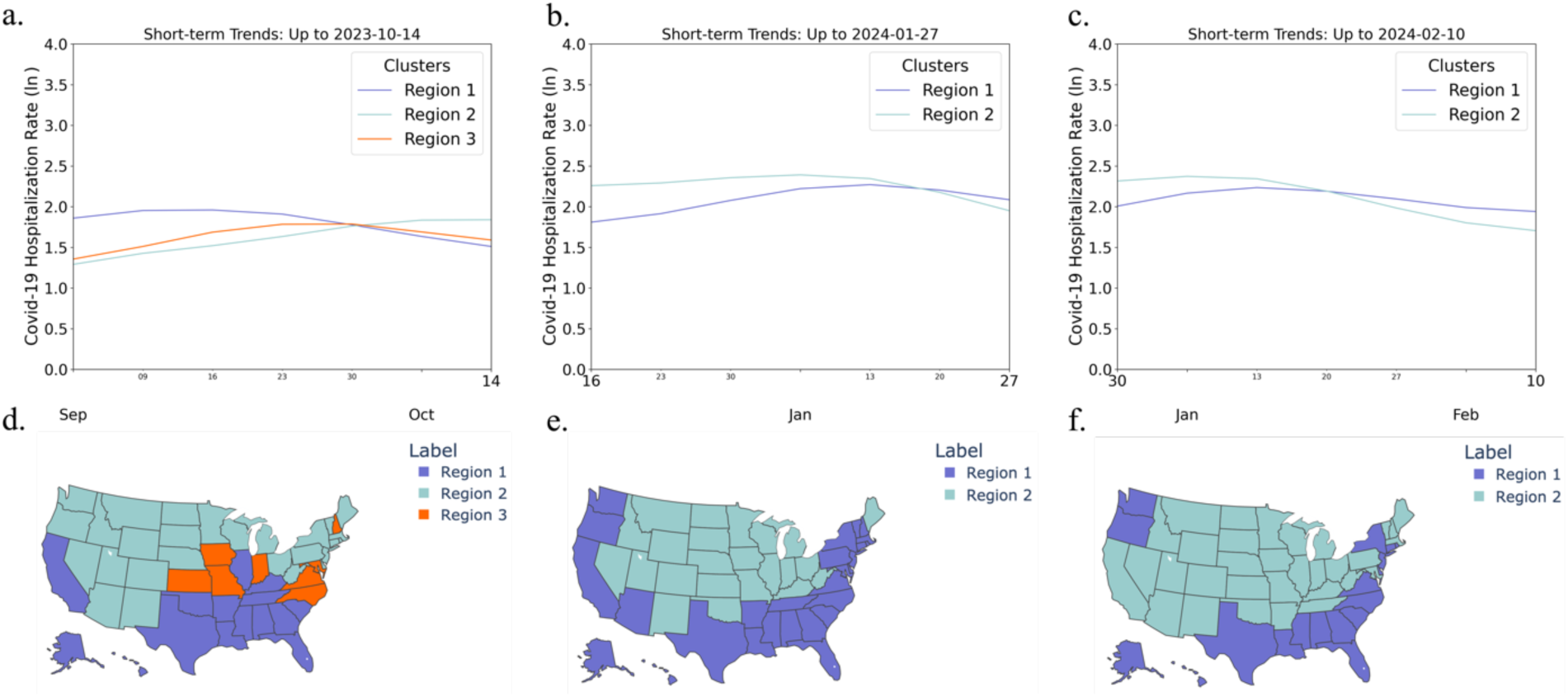
Short-term Hospitalization Trends Clustering for COVID-19 Hospitalization Rate in Selected Prediction Weeks: Subfigures (a, b, c) display the smoothed median influenza hospitalization rates for each clustered region (transformed using the natural logarithm, ln), while subfigures (d, e, f) show the corresponding states within those regions. The number of regions and the states within each region change dynamically based on short-term hospitalization trends.

Regions clustered based on long-term influenza hospitalization trends result in static groupings. Our clustering algorithms incorporate varying periods of historical data for both influenza and COVID-19. For influenza, we analyze hospitalization trends during the 2022-2023 season, as it represents the most recent ’normal’ influenza season after the disruptions caused by the COVID-19 pandemic. Figure S.7 below illustrates three distinct clusters, each showing significant differences in peak hospitalization rates and the timing of influenza prevalence.

**Figure S.7:**
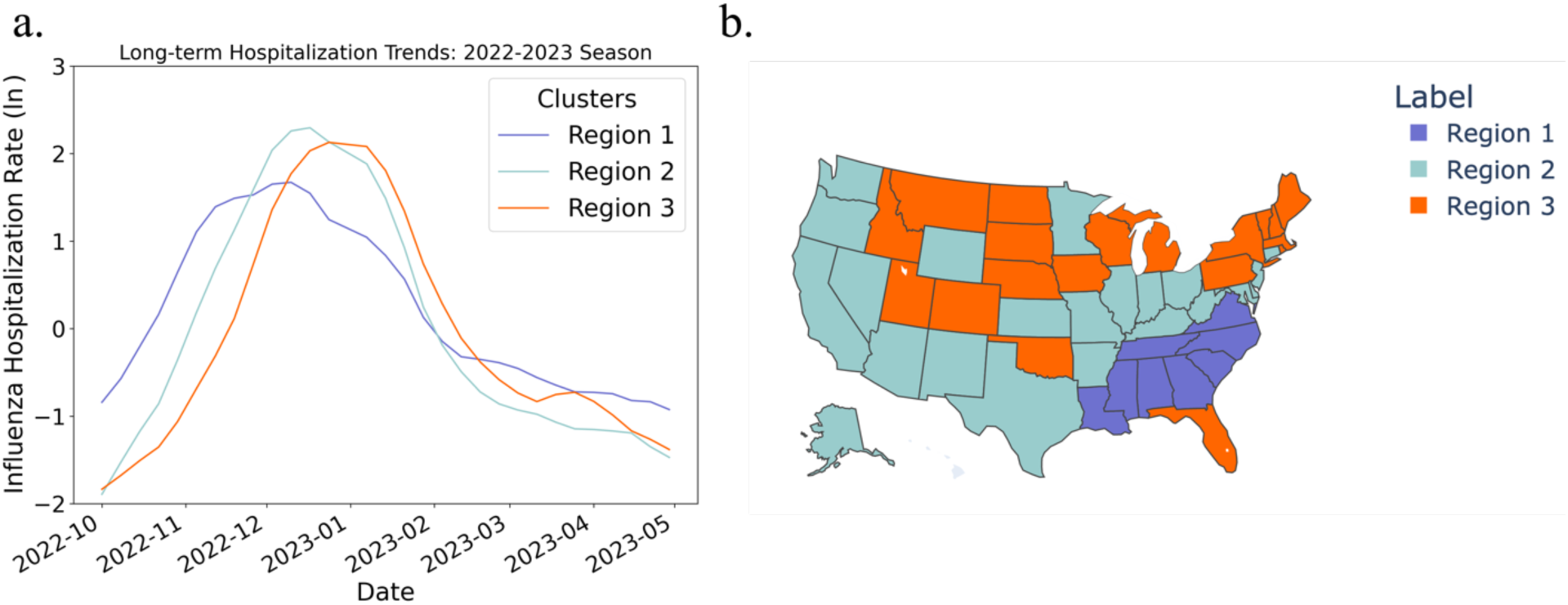
Long-term Hospitalization Trends Clustering for Influenza Hospitalization Rate (2022-2023 Influenza Season): Subfigure (a) display the smoothed median influenza hospitalization rates for each clustered region (transformed using the natural logarithm, ln), while subfigure (b) show the corresponding states within those regions.

For COVID-19, we cluster trends starting from the predominance of the Omicron variant through September 2023. This time frame was selected due to the significant impact Omicron had on transmission patterns and hospitalization rates, making it a key period for understanding the long-term dynamics of COVID-19. From Figure S.8, after May 2022, the peaks in COVID-19 admission rates differ between clusters, with one showing a distinct peak and the other displaying a more gradual, less defined pattern.

**Figure S.8:**
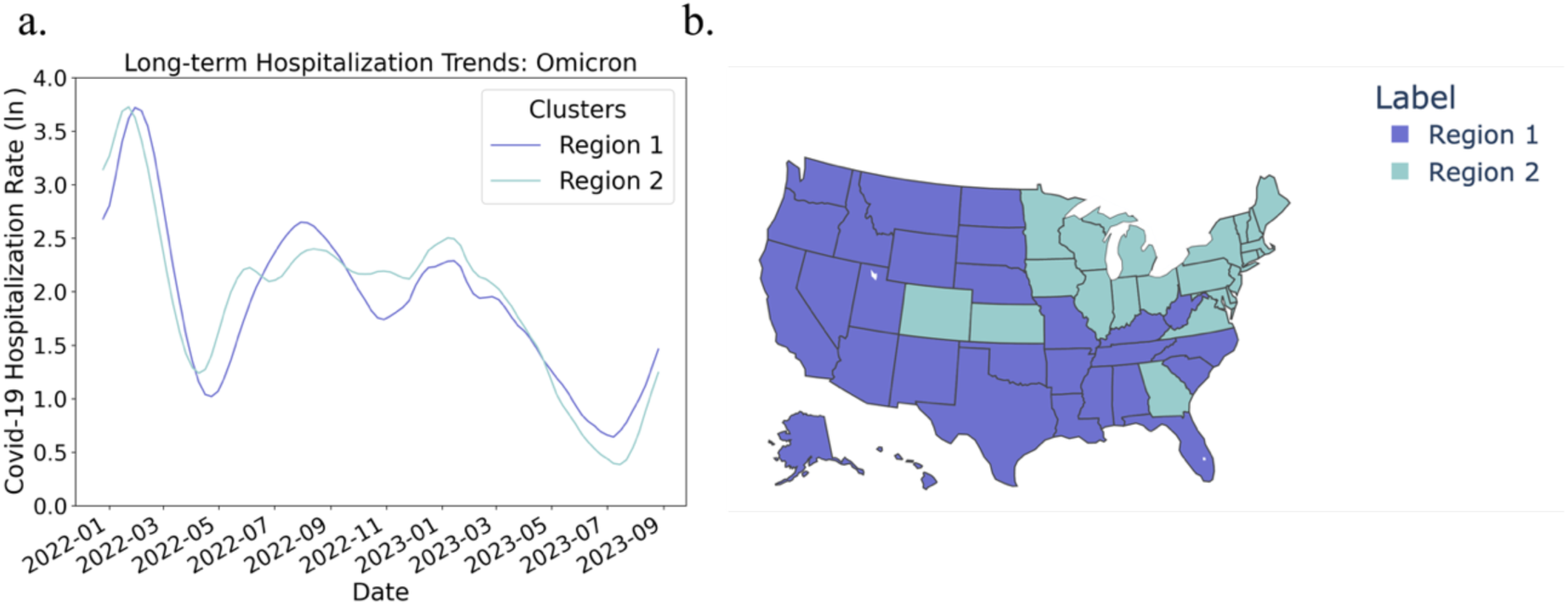
Long-term Hospitalization Trends Clustering for COVID-19 Hospitalization Rate (Omicron Wave): Subfigure (a) display the smoothed median COVID-19 hospitalization rates for each clustered region (transformed using the natural logarithm, ln), while subfigure (b) show the corresponding states within those regions.

The long-term clustering results for influenza reveal more pronounced distinctions between clusters compared to COVID-19, suggesting that influenza hospitalization patterns exhibit stronger regional variations.

Then, we evaluate the performance of regionally trained models for influenza and COVID-19 by comparing the MAE and WIS distributions. Figures S.9a and S.9c present the performance comparison of regionally trained models for influenza, while Figures S.9e and S.9g show the corresponding results for COVID-19. For both pathogens, regionally-trained models using data-driven clustering methods consistently outperform those based on predefined HHS regions, highlighting the effectiveness of our clustering approach.

For influenza, the regionally trained model using short-term trend clustering achieves the lowest MAE. It significantly outperforms the regionally-trained (HHS) model in both MAE and WIS, particularly at horizon 0, where it achieves more than an 18% improvement in the median and a 25% improvement in the upper quartile for MAE, along with a 16% improvement in the median and a 21% improvement in the upper quartile for WIS. For COVID-19, long-term trend clustering regions yield the best performance. It shows notable improvements over the HHS-based model, with a more than 21% reduction in median MAE and a 16% reduction in median WIS on average.

##### 3.3.2 Optimized Weighting Strategies

Next, we evaluate the performance gains achieved through ensemble weighting strategies, comparing the performance of the MEAN, RECI, RECI by horizon and LP approaches in aggregating the single-state model, the selected regionally-trained model, and the nationally trained model.

**Figure S.9:**
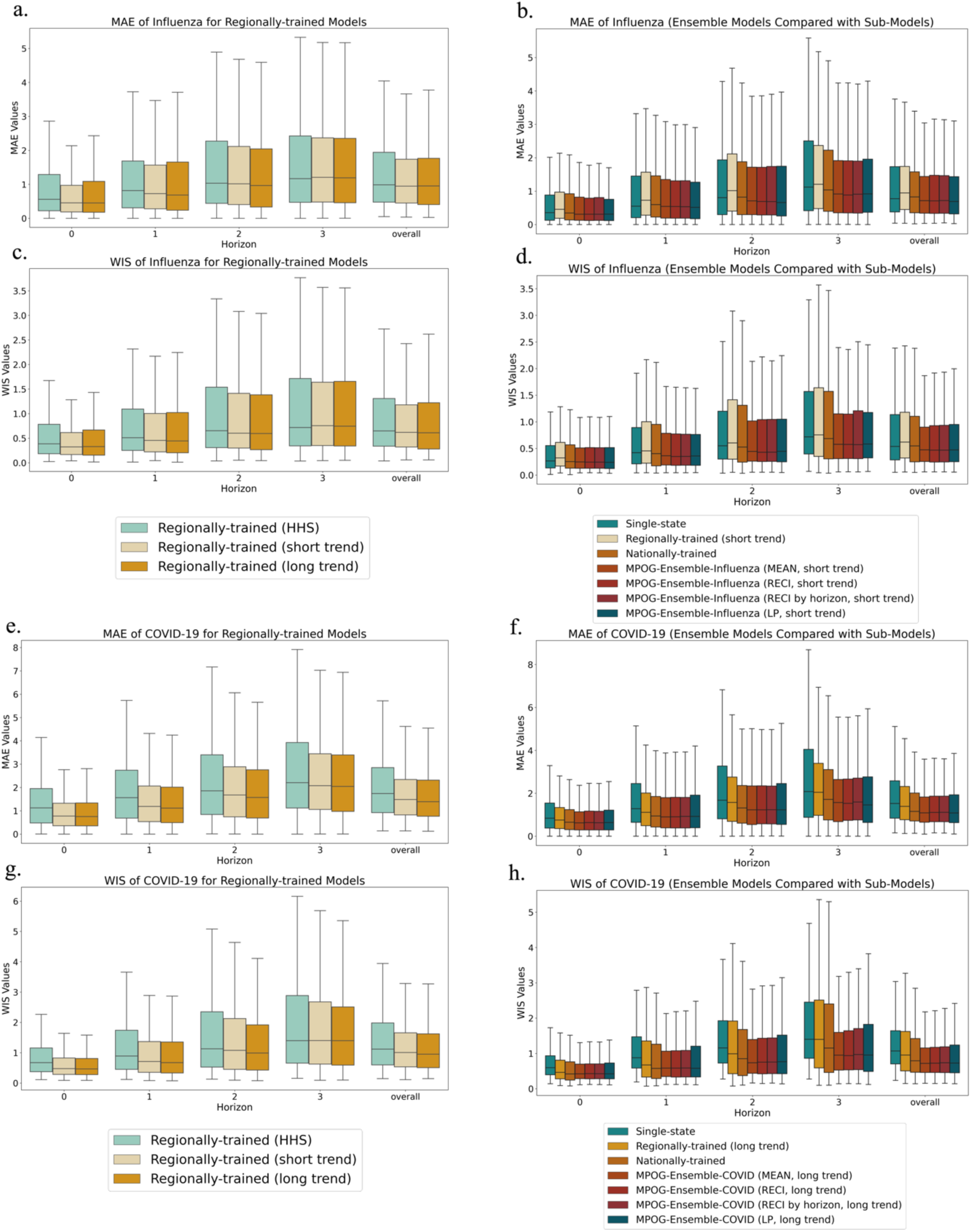
Box Plot of Errors for Regionally-trained Models (Left) and Ensemble Models Compared with Sub-Models (Right): Subfigures (a, b, c, d) present results for influenza models, while Subfigures (e, f, g, h) depict results for COVID-19 models. Each box in the plot represents the interquartile range (IQR) of the model errors. The interquartile range captures the middle 50% of the data, showing the spread of errors between the 25th percentile (lower edge) and the 75th percentile (upper edge). The line inside each box represents the median, which indicates the middle value of the error distribution. The whiskers extend to the smallest and largest values within 1.5 times the interquartile range from the box edges.

As illustrated in Figure S.9b and S.9d, the MPOG-Ensemble-Influenza models demonstrate over a 14% improvement compared to the Nationally-trained model and more than a 28% improvement over the regionally-trained (short trend) model in median MAE. For WIS, these improvements are 12% and 25%, respectively, highlighting the effectiveness of the ensemble approach. However, within each individual ensemble, the improvements are relatively modest, remaining below 2%, with MPOG-Ensemble-Influenza (LP, short trend) demonstrating the best performance.

For COVID-19 results shown in Figure S.9f and S.9h, the MPOG-Ensemble-COVID models achieve a 7% improvement in upper quartile MAE and a 16% improvement in upper quartile WIS compared to the nationally-trained model, with WIS improvements reaching up to 30% at horizon 3. These improvements highlight the ability of our ensemble methods to effectively reduce high error rates, particularly in the upper quartile, ensuring better performance across a range of scenarios.

The relatively small differences between each ensemble model can be attributed to the strong overall performance of all sub-models. However, when a poorly performing model, such as the FluSight Baseline or COVIDhub Baseline, is included in the ensemble, we observe significant differences across the ensemble methods. Figures S.10 and S.11 present box plots of WIS for the influenza and COVID-19 models, with the inclusion of a submodel of relatively lower performance. The results demonstrate that direct averaging is heavily impacted by the poor-performing model, resulting in significantly degraded performance. However, the RECI and RECI by horizon methods consistently assign lower weights to such models, effectively mitigating their negative impact. When the added model performs exceptionally poorly (e.g., the COVIDhub Baseline for COVID-19), the LP method proves most effective, optimizing weights based on past performance to produce a reliable forecast.

**Figure S.10:**
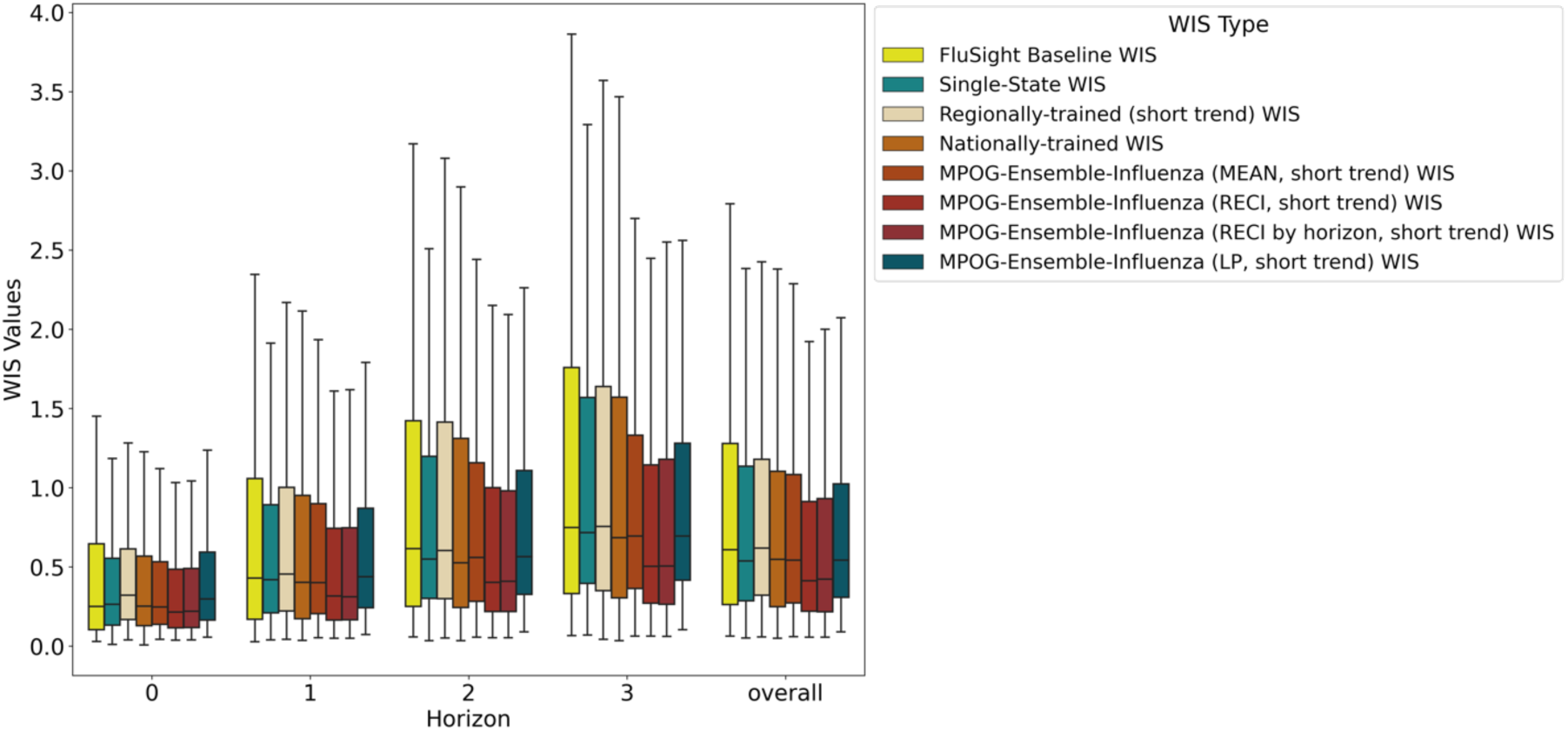
Box Plot of WIS (Influenza) with FluSight Baseline: Each box in the plot represents the interquartile range (IQR) of the model errors. The interquartile range captures the middle 50% of the data, showing the spread of errors between the 25th percentile (lower edge) and the 75th percentile (upper edge). The line inside each box represents the median, which indicates the middle value of the error distribution. The whiskers extend to the smallest and largest values within 1.5 times the interquartile range from the box edges.

**Figure S.11:**
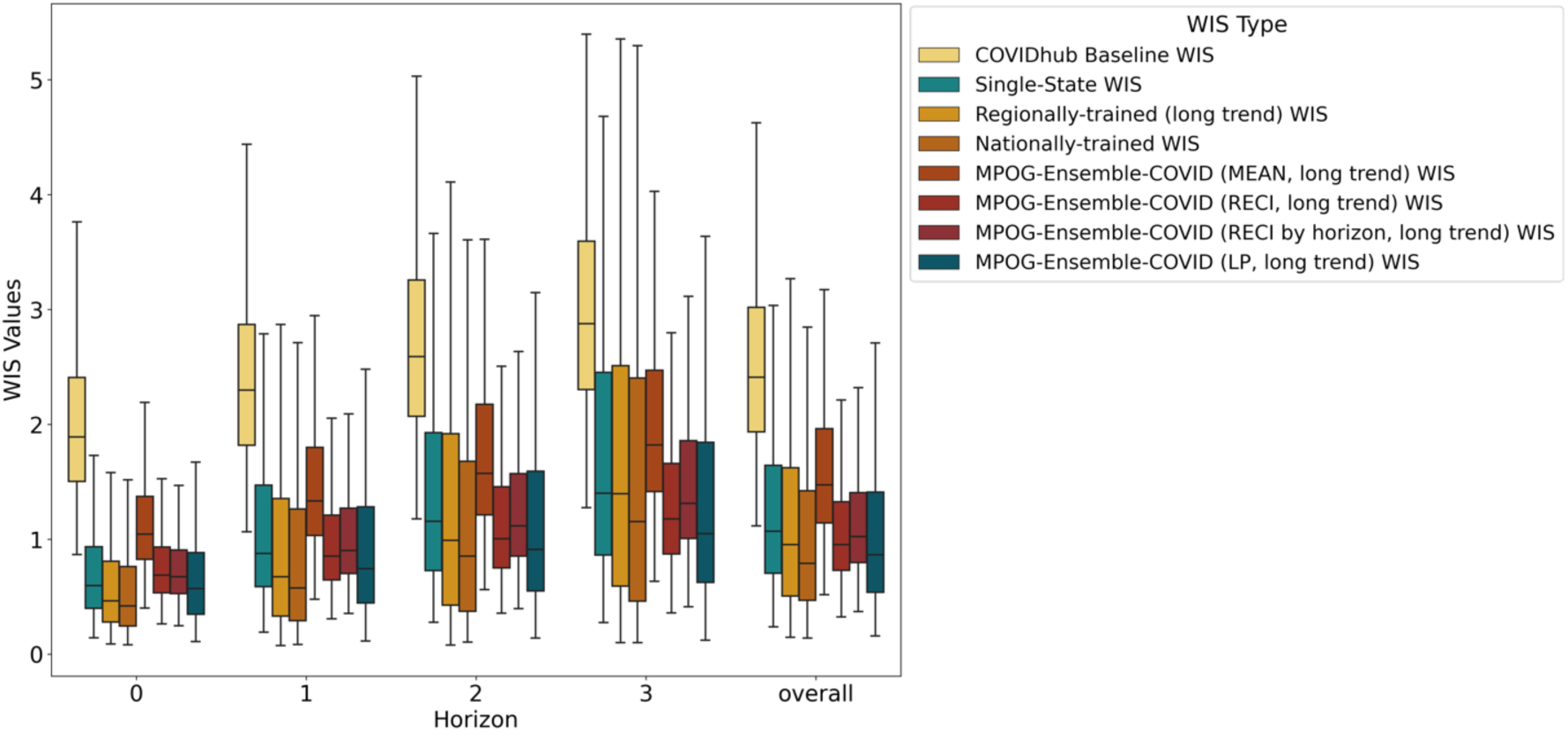
Box Plot of WIS (COVID-19) with COVIDhub Baseline: Each box in the plot represents the interquartile range (IQR) of the model errors. The interquartile range captures the middle 50% of the data, showing the spread of errors between the 25th percentile (lower edge) and the 75th percentile (upper edge). The line inside each box represents the median, which indicates the middle value of the error distribution. The whiskers extend to the smallest and largest values within 1.5 times the interquartile range from the box edges.

In cases where a sub-model performs poorly, simply averaging all models can decrease the ensemble performance significantly. In contrast, advanced methods such as RECI, RECI by horizon, and linear programming (LP) are more effective in preventing underperforming models from exerting undue influence on the ensemble. Specifically, when a model’s error is excessively large, LP assigns it a weight close to zero, minimizing its impact on the overall forecast. These findings demonstrate the robustness of our ensemble methods, effectively mitigating the influence of underperforming models and resulting in more reliable and accurate predictions.

##### 3.3.3 Dynamic Model Weights

This section presents how the weights assigned to the state, regionally-trained, and nationally-trained models change over time, as determined by the optimization methods. We showcase these weight changes specifically for the best-performing models for influenza and COVID-19.

Our ensemble methods exhibit similar patterns during key turning points for each pathogen. Specifically, the weight assigned to single-state models tends to decrease, with the linear programming (LP) method often reducing it to zero. In contrast, the nationally-trained and regionally-trained models assume greater importance, with the nationally-trained model frequently becoming dominant. Generally, regional models have a smaller impact on the ensemble, while the US-level model plays a more significant role in the forecasting process. In the following, we present a weight analysis of our best-performing models for influenza and COVID-19 forecasting.

The best-performing influenza MPOG-Ensemble model is weighted by linear programming and clustered based on short-term trends. Figure S.12 below demonstrates that when influenza hospitalization trends are stable, the single-state and Nationally-trained models dominate, while the regionally-trained model has minimal impact on the ensemble results. However, during turning points, the regionally-trained model becomes crucial. The MAE and WIS plots clearly show that during these turning points, particularly in December and January, our model significantly outperforms both the FluSight baseline and FluSight ensemble models, highlighting the effectiveness of our clustering and weighting strategy.

**Figure S.12:**
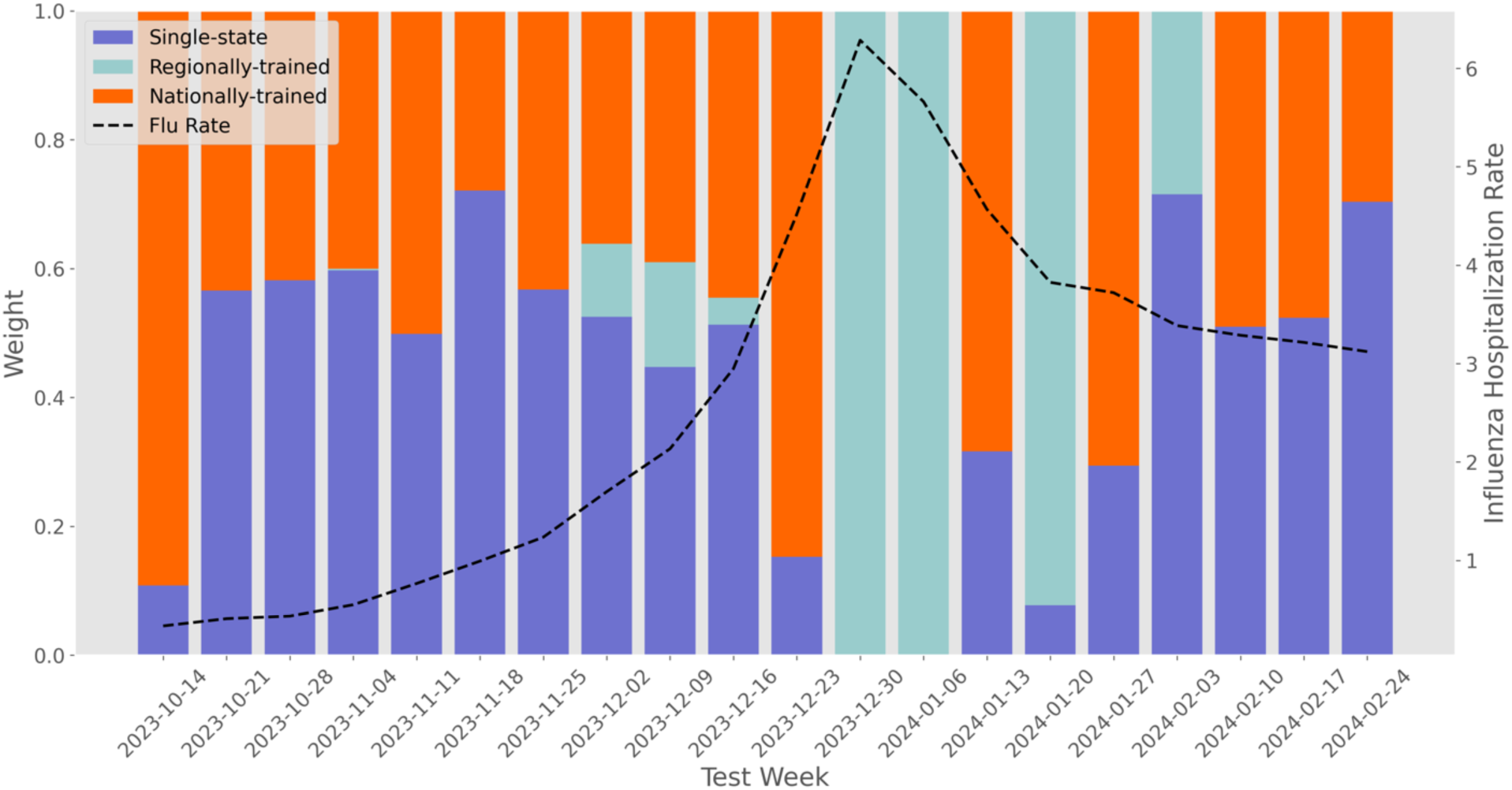
Weight Distribution Over Time for MPOG-Ensemble-Influenza (LP, short trend): The purple bar represents the weight assigned to the single-state model, the light blue bar corresponds to the regionally-trained model, and the orange bar indicates the weight for the national generalized model. The dashed black line shows the national influenza hospitalization rate over time.

The best-performing COVID-19 MPOG-Ensemble model is weighted using reciprocal weighting and based on long-term trends. Compared to Influenza model, the weights for COVID-19 are relative stable. As shown in Figure S.13, during turning points, the weight of the single-state model decreases slightly, while the weights of the US-level and regional models increase slightly.

**Figure S.13:**
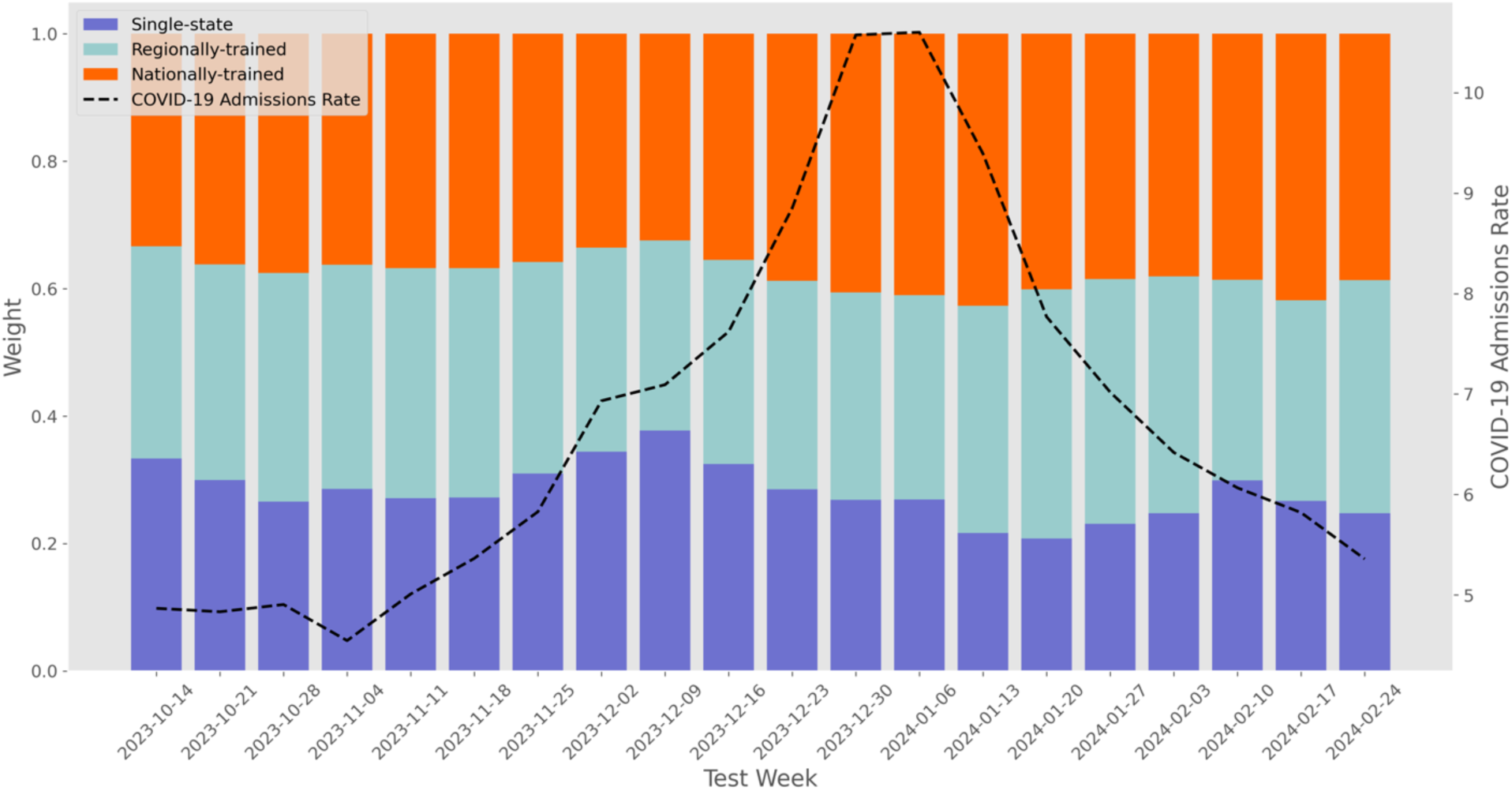
Weight Distribution Over Time for MPOG-Ensemble-COVID (RECI, long trend): The purple bar represents the weight assigned to the single-state model, the light blue bar corresponds to the regionally-trained model, and the orange bar indicates the weight for the national generalized model. The dashed black line shows the national COVID-19 hospitalization rate over time.

